# The clinically extremely vulnerable to COVID: Identification and changes in healthcare while self-isolating (shielding) during the coronavirus pandemic

**DOI:** 10.1101/2021.09.09.21263026

**Authors:** Jessica E. Butler, Mintu Nath, Dimitra Blana, William P. Ball, Nicola Beech, Corri Black, Graham Osler, Sebastien Peytrignet, Katie Wilde, Artur Wozniak, Simon Sawhney

## Abstract

**Background:** In March 2020, the government of Scotland identified people deemed clinically extremely vulnerable to COVID due to their pre-existing health conditions. These people were advised to strictly self-isolate (shield) at the start of the pandemic, except for necessary healthcare. We examined who was identified as clinically extremely vulnerable, how their healthcare changed during isolation, and whether this process exacerbated healthcare inequalities.

**Methods:** We linked those on the shielding register in NHS Grampian, a health authority in Scotland, to healthcare records from 2015-2020. We described the source of identification, demographics, and clinical history of the cohort. We measured changes in out-patient, in-patient, and emergency healthcare during isolation in the shielding population and compared to the general non-shielding population.

**Results:** The register included 16,092 people (3% of the population), clinically vulnerable primarily due to a respiratory disease, immunosuppression, or cancer. Among them, 42% were not identified by national healthcare record screening but added *ad hoc*, with these additions including more children and fewer economically-deprived.

During isolation, all forms of healthcare use decreased (25%-46%), with larger decreases in scheduled care than in emergency care. However, people shielding had better maintained scheduled care compared to the non-shielding general population: out-patient visits decreased 35% vs 49%; in-patient visits decreased 46% vs 81%. Notably, there was substantial variation in whose scheduled care was maintained during isolation: younger people and those with cancer had significantly higher visit rates, but there was no difference between sexes or socioeconomic levels.

**Conclusions:** Healthcare changed dramatically for the clinically extremely vulnerable population during the pandemic. The increased reliance on emergency care while isolating indicates that continuity of care for existing conditions was not optimal. However, compared to the general population, there was success in maintaining scheduled care, particularly in young people and those with cancer. We suggest that integrating demographic and primary care data would improve identification of the clinically vulnerable and could aid prioritising their care.

## INTRODUCTION

During the coronavirus pandemic of 2020, the Scottish Government and National Health Service Scotland sought to protect those who were most vulnerable to poor COVID outcomes by identifying people with serious underlying medical conditions and asking them to self-isolate (shield) at home^1–3^.

In Scotland, the self-isolation (shielding) period was initially 12 weeks of strict isolation from the 26^th^ March through to 18^th^ June 2020, which was extended with modification through 31^st^ July 2020 ^1,2^. People were asked to remain indoors as much as possible and to minimise interaction with others (including household members) to reduce the risk of contact with the virus ^2^. People advised to shield were eligible for furlough from work, statutory sick pay, and personal support such as home delivery of government-funded food and essential supplies ^3^.

The objective of the shielding programme was to protect clinically vulnerable people from the harm of getting COVID-19 by minimising avoidable care while maintaining the necessary care required to avoid harm from their underlying illnesses.

Correctly identifying those most vulnerable during the pandemic was critical – overlooking those at greatest risk could leave both the individual and the health system vulnerable to being overwhelmed by COVID and could prevent those in greatest need from accessing financial and social support. In contrast, asking people to isolate when they are not at substantial risk exposes them to physical and mental stress of isolation and potential consequences on education, careers, households and communities ^4,5^.

Even with perfect identification of the medically vulnerable, isolation at home for months could affect the medical care of people with existing chronic diseases. Healthcare systems sought to protect care with dedicated clinical facilities for shielding patients, but access may have been affected by changes in healthcare provider behaviour or in care-seeking behaviour of patients. Loss of care could have serious adverse consequences given the shielding population’s underlying chronic diseases, and had the potential to exacerbate existing age, social and demographic inequalities in access to care ^6–8^.

No population study has evaluated if maintenance of healthcare while shielding was achieved. Existing studies of healthcare use in the shielding population have shown a drop in healthcare use and a negative impact on health outcomes but have primarily covered specialised subsets of those on the shielding register and lack comparison with the non-shielding population^3,4,9–11^.

Here we present a study of the entire population of the shielding register in the NHS Grampian Health Board in Scotland (population 585,700). We describe the shielding population’s demographic and clinical profiles and evaluate differences in the methods used to identify individuals. We measure the shielded population’s use of healthcare services across all secondary care settings both before the first lockdown and during the first wave of the 2020 pandemic. We compare healthcare use before and during the lockdown between the shielding and non-shielding population in Grampian and for different sub-populations within the shielding population (by age, sex, and reason for shielding). We evaluate: (1) demographic differences in the vulnerable populations identified by different means; (2) whether people who shielded had changes in healthcare use; (3) if healthcare use changes in the shielding population were different to the rest of Grampian; (4) whether people who shielded for different underlying conditions had different changes in care use; and (5) whether these changes exacerbated existing social or demographic disparities in access to care.

## METHODS

### Identification of the clinically extremely vulnerable

In Scotland, people were formally recognised as clinically extremely vulnerable if their medical records showed they were in one of six categories: people with solid organ transplants, specific cancers, severe respiratory conditions, rare diseases, who were pregnant with significant heart disease, and who were on immunosuppression therapies ^12^.

People were identified as clinically extremely vulnerable by algorithmic searching of two primary datasets available at a national level – medications prescribed by GPs and diagnoses and procedure codes from in-hospital admissions ^12^. No other primary care records were available nationally and no sociodemographic characteristics were considered. However, general practitioners and hospital clinicians at local NHS boards were free to add people to the shielding register as they saw fit.

### Population

The study population was all people of all ages on the NHS Grampian shielding register who were alive at the start of shielding on 26^th^ March 2020. The study population’s healthcare use was compared to that of the rest of the Grampian population (total 585,700 and 569,608 non-shielding).

### Healthcare Use

Key information from the register of individuals advised to shield was provided by NHS Grampian included: the NHS identification number Community Health Index, date of addition to register, source of identification (locally by NHS Grampian or nationally by the Scottish Government), reasons for shielding, and date of death. We re-categorised people who were advised to shield due to being pregnant with a heart condition into the “other reasons” category for disclosure control due to their being fewer than 5 people. In cases where more than one reason for shielding was given for a person, a primary reason was defined with the following order of priority: cancer, transplant, respiratory disease, rare disease, immunosuppressants, other reason.

People on the shielding register were linked to their individual electronic healthcare records using their Community Health Index number. We extracted from the NHS Grampian TrakCare patient management system all admissions and attendances from 1^st^ January 2015 through 31^st^ July 2020 at the following facilities: accident and emergency, hospital in-patient (including children’s hospital), and out-patient clinics with any specialty. All accident and emergency attendances were categorised as emergency visits and all out-patient attendances were categorised as scheduled visits. In-patient admissions could be either emergency or scheduled – this status of the admission was taken from NHS Scotland Information Services Division General Acute Inpatient and Day Case - Scottish Morbidity Record (SMR01)^13^. ICD-10 codes ^14^ for main and contributory diagnoses during in-patient admissions were also taken from Scottish Morbidity Record SMR01. Morbidity counts and weights were calculated using all ICD-10 codes associated with in-patient admissions from the five-year period of 1^st^ March 2015 to 1^st^ March 2020 using the R package comorbidity ^15^ using Elixhauser morbidities ^16^ and van Walraven weighted score ^17^. Home area demographics were taken from the data zone of the home postcode given in the Community Health Index database. Home area deprivation (a combined relative measure of income, employment, education, health, access to services, crime and housing) was taken from the Scottish Government’s Index of Multiple Deprivation (SIMD2020v2)^18^ and urban-rural classification (with these categories: Large Urban Areas, Other Urban Areas, Accessible Small Towns, Remote Small Towns, Accessible Rural Areas, Remote Rural Areas) was taken from Scottish Government Urban Rural classification for 2016^19^. Areas of deprivation were defined as those in the most deprived 20% of Scotland according to SIMD.

For healthcare use by the Grampian population as a whole, NHS Grampian provided total counts across the entire population per day for: all emergency department attendances, in-patient admissions, and out-patient clinic attendances from their TrakCare patient management system.

### Statistical analysis

#### Time periods

Healthcare use for both shielding and general non-shielding population was modelled at two time periods (phases): the pre-shielding period between 1^st^ January and 14^th^ March 2020 and the shielding period between 22^nd^ March and 18^th^ June 2020 which marked the end of the initial national shielding period. We excluded the data during the two weeks of the transition period (between 15th and 21st of March).

#### Comparison of healthcare use between those shielding vs the rest of the Grampian population

Total weekly healthcare use between these two phases was compared between the shielding population and the rest of the Grampian population. We fitted separate generalised linear models for each healthcare type (emergency, emergency in-patient, scheduled in-patient, and out-patient) and adopted robust model selection strategies. We observed a negative binomial distribution of the healthcare data performed best^20^. The model included a logarithmic link function and the logarithm of respective population size as an offset variable.

#### Healthcare use of patients on the shielding register

Data from patients on the shielding register were further analysed to evaluate whether patient characteristics (age, sex, home-area deprivation and primary reason for shielding) were associated with the changes in healthcare use during the shielding period. First, we categorised these characteristics as follows: age (5 levels: <50, 50-59, 60-69, 70-79, 80+ years); deprivation into two levels using Scottish Index of Multiple Deprivation (SIMD) decile rank scores: not-deprived (SIMD2020 version 2 decile > 2) and deprived (SIMD2020 version 2 decile <= 2); and dominant shielding reason (six levels in the following order of priority: cancer, transplant, respiratory, rare, immunosuppressants and other). For patients with two or more shielding reasons, the top priority reason was assigned. We then summarised and created patient groups for the levels of sex, age, deprivation and shielding reasons and calculated total counts of healthcare use at each phase. The summarised grouped data at each phase (120 combinations) are referred to here as a “group”. The length of stay at each phase for each group was calculated as the median length of stay for all patients for the corresponding group and phase combination. Similarly, accounting for the mortality records, the total number of surviving patients for each group and phase combination was calculated.

We fitted a generalised linear mixed model on the summarised data of each healthcare use. As noted earlier, a negative binomial distribution of the outcome variable showed the best performance. The model considered the logarithmic link function and included age, sex, deprivation, shielding reasons and phase of lockdown as fixed effects, and group as a random effect. The model also included the logarithm of median length of stay and the logarithm of the number of surviving patients as offset variables. We hypothesised that changes in healthcare use may vary between sex, deprivation and shielding reasons at both phases. Therefore, we assessed reasonable two and three-way interaction terms using a Wald chi-squared test and the final model included statistically significant interaction terms (p<0.05). An intermediate model for each outcome variable is also presented to investigate the two-way interaction effect of sex, deprivation and shielding group with phase. All statistical models were fitted in the R software environment using libraries mgcv^21^ and glmmTB^22^.

All code used to complete the above data processing and statistical analysis is given in Supplementary Files.

### Project approvals and information governance

This project was approved by the North Node Privacy Advisory Committee (NNPAC) (project ID: 6-081-20). NNPAC provides researchers with streamlined access to NHS Grampian data for research purposes, and committee approval incorporates approvals from: project sponsor, ethics panel, the Caldicott Guardian, and NHS R&D. All analysis was carried out in the Grampian Data Safe Haven (project ID: DaSH412) on pseudonomysed individual-level data. Per UK General Data Protection Regulation, only aggregate data can be released from the Grampian Data Safe Haven for publication, but all individual-level data has been archived and can be accessed by application to the Grampian Data Safe Haven.

## RESULTS

### Identification of the clinically extremely vulnerable for the shielding register

Table 1 describes the sociodemographic and key clinical characteristics of people on the shielding register. 16,092 of 585,700 people in NHS Grampian (2.7%) were identified as clinically extremely vulnerable to COVID and placed on the shielding register. The most common underlying health condition necessitating shielding was chronic respiratory disease (41% of the shielding population), followed by immunosuppression (21%) and active cancer treatment (18%) (Table 1).

**Table 1.** Demographics of the shielding register population in the NHS Grampian region. Demographics of the shielding population in total and stratified by: source of identification (nationally via an electronic health record scan done by the Scottish Government or *ad hoc* locally by GPs and hospital clinicians in NHS Grampian); primary underlying health condition requiring shielding; sex; age group; Scottish Index of Multiple Deprivation decile for area of residence; Scottish Government Urban Rural Classification of area of residence.

|  | N | Women | Age (med) | People <20 years old | Deprived Home Area | Remote or Rural | Identified Nationally |
| --- | --- | --- | --- | --- | --- | --- | --- |
| **TOTAL SHIELDING REGISTER** |  |  |  |  |  |  |  |
|  | 16092 | 8632 (54%) | 65 | 496 (3%) | 1230 (8%) | 5494 (34%) | 9354 (58%) |
| **SOURCE OF IDENTIFICATION** |  |  |  |  |  |  |  |
| national health record scan | 9354 | 4975 (53%) | 66 | 218 (2%) | 792 (8%) | 3103 (33%) | 9354 (100%) |
| locally *ad hoc* by clinicians | 6738 | 3657 (54%) | 63 | 278 (4%) | 438 (7%) | 2391 (35%) | 0 (0%) |
| **SHIELDING REASON** |  |  |  |  |  |  |  |
| respiratory disease | 6535 | 3565 (55%) | 68 | 136 (2%) | 657 (10%) | 2167 (33%) | 5566 (85%) |
| immunosuppressants | 3437 | 2089 (61%) | 57 | 149 (4%) | 189 (5%) | 1245 (36%) | 1469 (43%) |
| cancer | 2834 | 1476 (52%) | 68 | 55 (2%) | 157 (6%) | 1007 (36%) | 704 (25%) |
| other | 1783 | 807 (45%) | 62 | 66 (4%) | 130 (7%) | 587 (33%) | 477 (27%) |
| rare disease | 937 | 450 (48%) | 62 | 65 (7%) | 69 (7%) | 300 (32%) | 588 (63%) |
| transplant | 566 | 245 (43%) | 53 | 25 (4%) | 28 (5%) | 188 (33%) | 550 (97%) |
| **SEX** |  |  |  |  |  |  |  |
| female | 8632 | 8632 (100%) | 64 | 217 (3%) | 657 (8%) | 2880 (33%) | 4975 (58%) |
| male | 7460 | 0 (0%) | 65 | 279 (4%) | 573 (8%) | 2614 (35%) | 4379 (59%) |
| **AGE** |  |  |  |  |  |  |  |
| 0-19 | 496 | 217 (44%) | 11 | 496 (100%) | 37 (7%) | 147 (30%) | 218 (44%) |
| 20-29 | 530 | 288 (54%) | 25 |  | 33 (6%) | 153 (29%) | 325 (61%) |
| 30-39 | 905 | 484 (53%) | 35 |  | 77 (9%) | 256 (28%) | 480 (53%) |
| 40-49 | 1440 | 825 (57%) | 45 |  | 132 (9%) | 466 (32%) | 750 (52%) |
| 50-59 | 2666 | 1539 (58%) | 55 |  | 232 (9%) | 901 (34%) | 1437 (54%) |
| 60-69 | 3869 | 2076 (54%) | 65 |  | 303 (8%) | 1357 (35%) | 2330 (60%) |
| 70-79 | 4042 | 2086 (52%) | 74 |  | 274 (7%) | 1475 (36%) | 2455 (61%) |
| 80-89 | 1928 | 999 (52%) | 83 |  | 131 (7%) | 663 (34%) | 1232 (64%) |
| 90+ | 216 | 118 (55%) | 91 |  | 11 (5%) | 76 (35%) | 127 (59%) |
| **HOME AREA DEPRIVATION** |  |  |  |  |  |  |  |
| 1 (most deprived) | 229 | 113 (49%) | 61 | 3 (1%) | 229 (100%) | 0 (0%) | 151 (66%) |
| 2 | 1001 | 544 (54%) | 64 | 34 (3%) | 1001 (100%) | 0 (0%) | 641 (64%) |
| 3 | 1347 | 745 (55%) | 63 | 44 (3%) |  | 188 (14%) | 869 (65%) |
| 4 | 1727 | 928 (54%) | 64 | 39 (2%) |  | 395 (23%) | 1086 (63%) |
| 5 | 1554 | 877 (56%) | 65 | 55 (4%) |  | 721 (46%) | 968 (62%) |
| 6 | 2153 | 1153 (54%) | 66 | 62 (3%) |  | 1207 (56%) | 1253 (58%) |
| 7 | 2042 | 1103 (54%) | 66 | 49 (2%) |  | 1043 (51%) | 1216 (60%) |
| 8 | 2328 | 1203 (52%) | 65 | 80 (3%) |  | 1374 (59%) | 1269 (55%) |
| 9 | 1765 | 959 (54%) | 66 | 60 (3%) |  | 516 (29%) | 920 (52%) |
| 10 (least deprived) | 1766 | 917 (52%) | 65 | 54 (3%) |  | 50 (3%) | 879 (50%) |
| **HOME AREA RURALITY** |  |  |  |  |  |  |  |
| Accessible Rural | 3202 | 1656 (52%) | 65 | 86 (3%) | 0 (0%) | 3202 (100%) | 1763 (55%) |
| Accessible Towns | 1434 | 767 (53%) | 64 | 55 (4%) | 0 (0%) |  | 801 (56%) |
| Large Urban Areas | 5907 | 3224 (55%) | 64 | 184 (3%) | 886 (15%) |  | 3540 (60%) |
| Other Urban Areas | 3077 | 1671 (54%) | 66 | 94 (3%) | 344 (11%) |  | 1808 (59%) |
| Remote Rural | 1268 | 653 (51%) | 67 | 40 (3%) | 0 (0%) | 1268 (100%) | 680 (54%) |
| Remote Towns | 1024 | 571 (56%) | 67 | 21 (2%) | 0 (0%) | 1024 (100%) | 660 (64%) |

58% of the total shielding register population identified by the national health record scan and 42% was added *ad hoc* by clinicians. There was variation in whether people were *ad hoc* by underlying disease, age and home area deprivation. 15% of respiratory patients were not identified in the national analysis and were added *ad hoc*, compared to 75% of cancer patients (Table 1 & Figure 1A). 66% of children on the register were added *ad hoc*, compared to 41% of adults (Table 1 & Figure 1B). The proportion of *ad hoc* inclusion decreased with increasing home area deprivation (Table 1 & Figure 1C). Compared to several other sites in the UK, the Grampian shielding register had more people (6,738 or 42%) who were added by local experts^23^.

**Figure 1.**
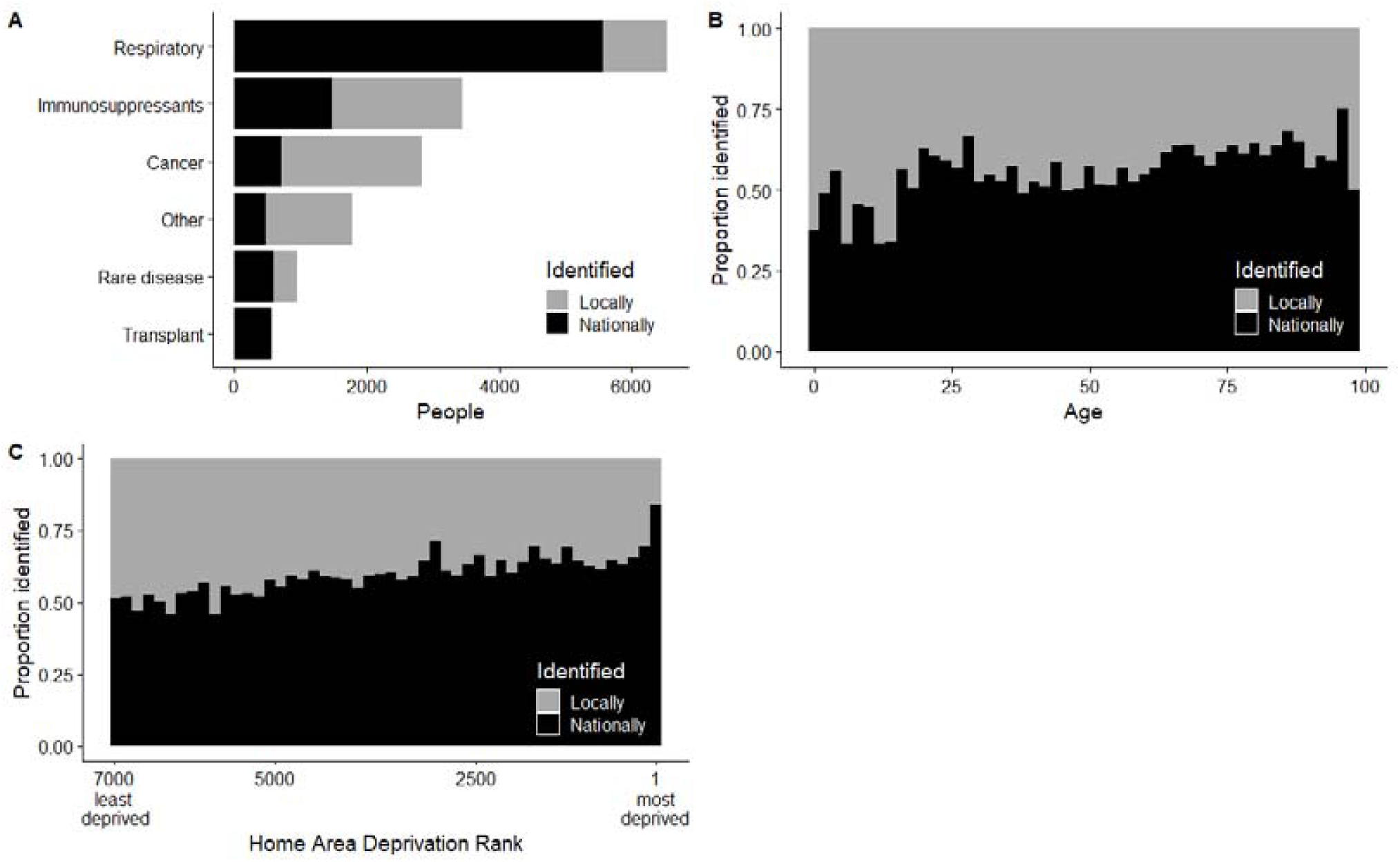
Demographic and clinical characteristics of people identified as clinically extremely vulnerable by national electronic health record scan by the Scottish Government versus by local analysis by GPs and hospital clinicians in NHS Grampian. A. Number of people identified nationally vs locally by primary underlying health condition requiring shielding, B. Proportion identified nationally vs locally by age, C. Proportion identified nationally vs locally by Scottish Index of Multiple Deprivation rank for area of residence

The shielding population was demographically diverse across the different clinical reasons for shielding. The three largest clinical populations (those with respiratory disease, on immunosuppressants, or with cancer) all had larger proportions of women (Table 1 and Figure 2A). Age varied from a median of 53 years among transplant patients to 68 in those with respiratory disease (Table 1 and Figure 2B). Proportionately more people living in the most deprived areas of Grampian were shielded due to respiratory disease and fewer due to immunosuppressants or cancer (Table1 and Figure 2C).

**Figure 2.**
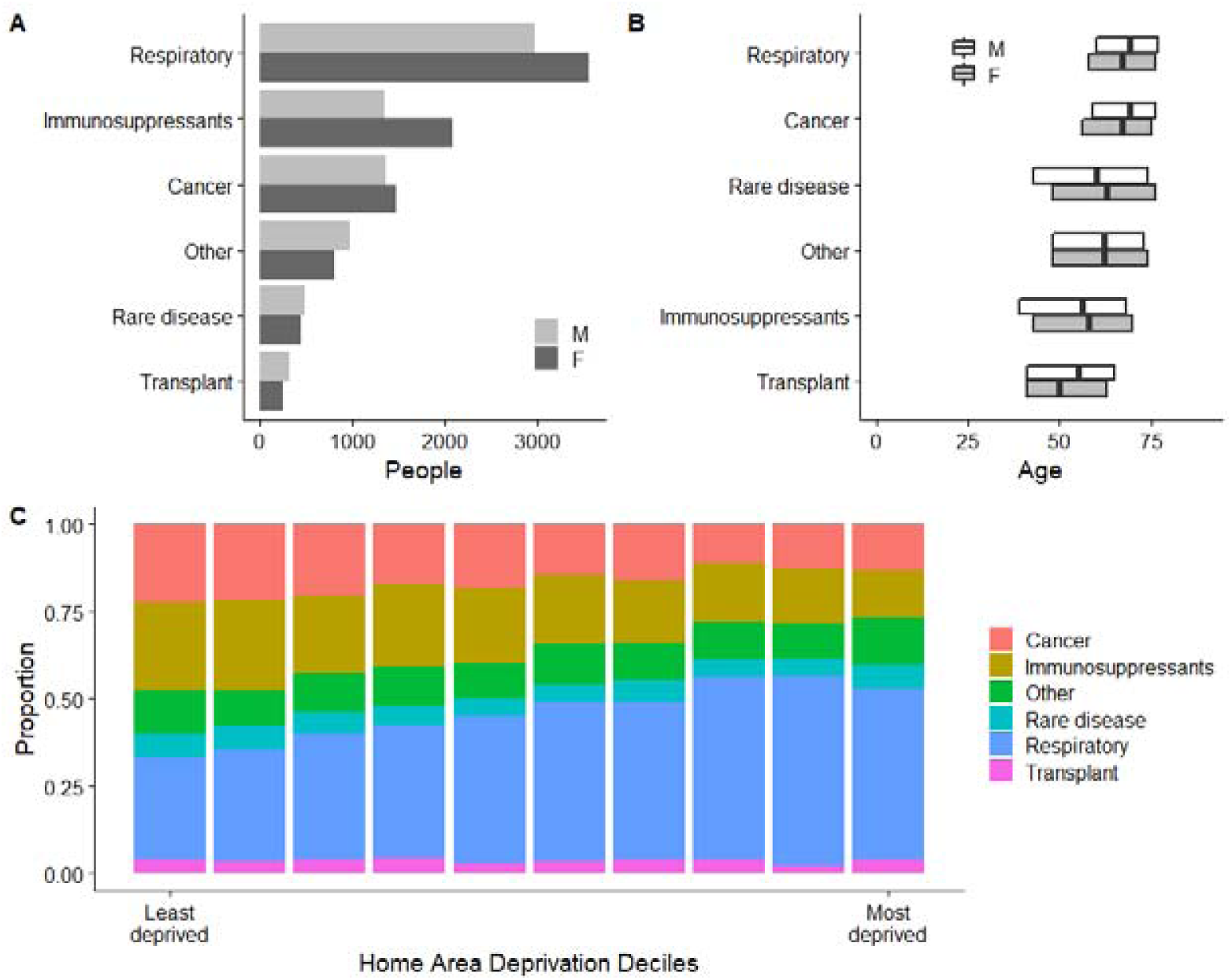
Demographic characteristics of the six primary clinical reasons for shielding register membership. A. Number of people by sex, B. Age (showing median and interquartile range) by sex, C. Proportion of population per decile of the Scottish Index of Multiple Deprivation decile for area of residence

### Pre-pandemic healthcare use by the shielding register population

Table 2 reports the healthcare during the one-year pre-pandemic (1^st^ March 2019 through 28^th^ February 2020) for the total shielding register and for key sub-populations. 84% of shielding patients had at least one out-patient attendance, 28% had a scheduled in-patient admission, 24% had an emergency in-patient admission, and 30% had an A&E attendance (Table 2). Shielding patients had a mean of 8 scheduled care attendances (five times more than the mean for the non-shielding population) and a mean of 1 emergency care attendance (3 times higher than the non-shielding population average) (Table 2). Overall, the shielding population made up 3% of the total population, but in the year pre-pandemic, had 14% of the scheduled care and 9% of the emergency care in Grampian – a total of 131,000 scheduled visits and 16,000 emergency visits (Table 2).

**Table 2.** Pre-pandemic annual healthcare use in the shielding register population. Healthcare use from 1^st^ March 2019 to 28^th^ February 2020 for the shielding population total and by source of identification, primary underlying health condition requiring shielding, sex, age, Scottish Index of Multiple Deprivation decile for area of residence, Scottish Government Urban Rural Classification of area of residence. OP– outpatient attendances, IP scheduled–scheduled inpatient admission, IP emergency–emergency inpatient admission, A&E–accident and emergency attendance, scheduled visits-all scheduled care in out-patient and in-patient settings, emergency visits–all unscheduled care in in-patient and A&E settings, multimorbidity–presence of more than one chronic disease diagnosis^24^ from inpatient admission in the previous 5 years, morbidity burden–weighted sum of chronic disease diagnoses^25^ from inpatient admission in previous 5 years.

|  | N | % with OP | % with IP sched | % with IP emerg | % with A&E | Sched visits | Emerg visits | % Visits emerg | Sched visits (mean/ person) | Emerg visits (mean/ person) | % Multi-morbid | Morbid. Burden (med) |
| --- | --- | --- | --- | --- | --- | --- | --- | --- | --- | --- | --- | --- |
| **TOTAL SHIELDING REGISTER** | 16,092 | 84% | 28% | 24% | 30% | 131,431 | 15,973 | 11% | 8.2 | 1 | 38% | 5.6 |
| **SOURCE OF IDENTIFICATION** |  |  |  |  |  |  |  |  |  |  |  |  |
| national health record scan | 9,354 | 79% | 24% | 23% | 30% | 61,400 | 9,150 | 13% | 6.6 | 1 | 40% | 5.5 |
| locally *ad hoc* by clinicians | 6,738 | 91% | 32% | 26% | 30% | 70,031 | 6,823 | 9% | 10.4 | 1 | 36% | 5.9 |
| **SHIELDING REASON** |  |  |  |  |  |  |  |  |  |  |  |  |
| respiratory disease | 6,535 | 75% | 21% | 24% | 31% | 36,956 | 6,798 | 16% | 5.7 | 1 | 43% | 5.3 |
| immunosuppressants | 3,437 | 91% | 25% | 16% | 22% | 25,293 | 2,037 | 7% | 7.4 | 0.6 | 23% | 2.4 |
| cancer | 2,834 | 93% | 48% | 35% | 38% | 43,314 | 4,016 | 8% | 15.3 | 1.4 | 45% | 8.7 |
| other | 1,783 | 83% | 25% | 26% | 31% | 13,755 | 1,812 | 12% | 7.7 | 1 | 40% | 6.3 |
| rare disease | 937 | 85% | 22% | 19% | 26% | 5,788 | 664 | 10% | 6.2 | 0.7 | 32% | 3.7 |
| transplant | 566 | 93% | 33% | 31% | 30% | 6,325 | 646 | 9% | 11.2 | 1.1 | 65% | 10 |
| **SEX** |  |  |  |  |  |  |  |  |  |  |  |  |
| female | 8,632 | 85% | 27% | 24% | 30% | 71,058 | 8,491 | 11% | 8.2 | 1 | 37% | 5.3 |
| male | 7,460 | 82% | 28% | 25% | 29% | 60,373 | 7,482 | 11% | 8.1 | 1 | 40% | 6 |
| **AGE** |  |  |  |  |  |  |  |  |  |  |  |  |
| 0-19 | 496 | 92% | 39% | 39% | 28% | 7,063 | 901 | 11% | 14.2 | 1.8 | 16% | 2.9 |
| 20-29 | 530 | 85% | 20% | 20% | 30% | 3,796 | 537 | 12% | 7.2 | 1 | 15% | 2.9 |
| 30-39 | 905 | 84% | 23% | 18% | 27% | 7,356 | 792 | 10% | 8.1 | 0.9 | 17% | 3.4 |
| 40-49 | 1,440 | 83% | 21% | 20% | 26% | 10,742 | 1,231 | 10% | 7.5 | 0.9 | 22% | 3.9 |
| 50-59 | 2,666 | 82% | 25% | 20% | 27% | 21,163 | 2,246 | 10% | 7.9 | 0.8 | 29% | 4.8 |
| 60-69 | 3,869 | 82% | 29% | 22% | 28% | 31,251 | 3,506 | 10% | 8.1 | 0.9 | 39% | 5.6 |
| 70-79 | 4,042 | 85% | 32% | 26% | 31% | 34,262 | 4,006 | 10% | 8.5 | 1 | 50% | 6.6 |
| 80-89 | 1,928 | 86% | 27% | 33% | 38% | 14,704 | 2,400 | 14% | 7.6 | 1.2 | 59% | 7.6 |
| 90+ | 216 | 75% | 16% | 41% | 48% | 1,094 | 354 | 24% | 5.1 | 1.6 | 57% | 7.6 |
| **HOME AREA DEPRIVATION** |  |  |  |  |  |  |  |  |  |  |  |  |
| 1 (most deprived) | 229 | 83% | 23% | 26% | 37% | 1,593 | 294 | 16% | 7 | 1.3 | 41% | 4.8 |
| 2 | 1,001 | 83% | 27% | 28% | 38% | 7,900 | 1,353 | 15% | 7.9 | 1.4 | 45% | 5.7 |
| 3 | 1,347 | 84% | 28% | 27% | 33% | 10,764 | 1,559 | 13% | 8 | 1.2 | 43% | 5.6 |
| 4 | 1,727 | 84% | 26% | 25% | 29% | 14,141 | 1,693 | 11% | 8.2 | 1 | 41% | 5.8 |
| 5 | 1,554 | 82% | 28% | 23% | 31% | 11,700 | 1,472 | 11% | 7.5 | 0.9 | 37% | 5.4 |
| 6 | 2,153 | 82% | 28% | 25% | 30% | 17,355 | 2,173 | 11% | 8.1 | 1 | 41% | 5.7 |
| 7 | 2,042 | 84% | 28% | 24% | 29% | 15,554 | 1,974 | 11% | 7.6 | 1 | 39% | 5.6 |
| 8 | 2,328 | 84% | 28% | 24% | 28% | 19,666 | 2,164 | 10% | 8.4 | 0.9 | 35% | 5.7 |
| 9 | 1,765 | 87% | 27% | 23% | 27% | 15,047 | 1,521 | 9% | 8.5 | 0.9 | 34% | 5.7 |
| 10 (least deprived) | 1,766 | 87% | 29% | 24% | 30% | 17,196 | 1,699 | 9% | 9.7 | 1 | 37% | 5.6 |
| **HOME AREA RURALITY** |  |  |  |  |  |  |  |  |  |  |  |  |
| Accessible Rural | 5,907 | 85% | 27% | 26% | 34% | 54,256 | 6,682 | 11% | 9.2 | 1.1 | 40% | 5.7 |
| Accessible Towns | 3,077 | 84% | 29% | 25% | 30% | 23,040 | 3,172 | 12% | 7.5 | 1 | 39% | 5.8 |
| Large Urban Areas | 1,434 | 84% | 28% | 24% | 29% | 12,228 | 1,357 | 10% | 8.5 | 0.9 | 37% | 5.8 |
| Other Urban Areas | 3,202 | 83% | 27% | 23% | 27% | 24,531 | 2,815 | 10% | 7.7 | 0.9 | 36% | 5.5 |
| Remote Rural | 1,024 | 83% | 28% | 22% | 26% | 7,115 | 790 | 10% | 6.9 | 0.8 | 41% | 5.3 |
| Remote Towns | 1,268 | 84% | 30% | 23% | 25% | 9,746 | 1,086 | 10% | 7.7 | 0.9 | 39% | 5.7 |

Patterns of healthcare use varied widely across demographics and reasons for shielding. Cancer patients used the most care, with an average of 15 scheduled visits and 1.5 emergency visits in the year pre-pandemic (Table 2). Respiratory patients had less scheduled care (6 visits/year) but required proportionally more emergency care (16% of their care was emergency, twice the rate of cancer patients) (Table 2).

People aged under 20 years had the highest levels of both scheduled care (14 visits/year) and emergency care (2 visits/year) (Table 2 & Figure 3). Scheduled care was substantially lower for people over aged 70 (5 visits/year), whereas emergency care substantially was higher in this elderly subset (Table 2).

**Figure 3.**
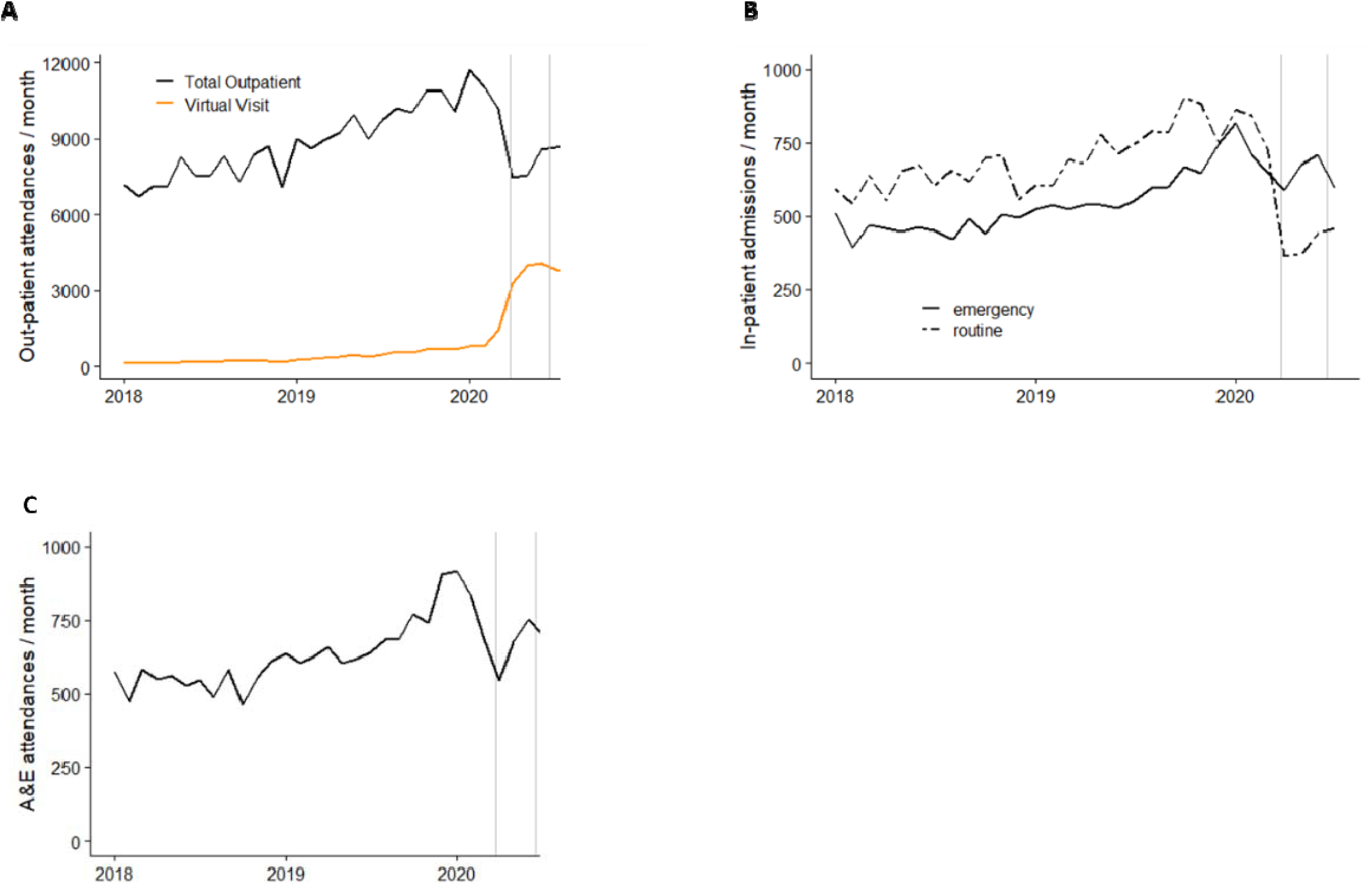
Monthly healthcare use by the shielding register population. Attendances and admissions per month for the shielding population. A) Out-patient attendances per month (total and virtual), b) in-patient admissions per month (scheduled and emergency), and C) emergency attendances per month. Gray lines show the initial shielding period 26th March to 18th June 2020.

People living in the most deprived areas had fewer scheduled healthcare visits compared to the least deprived (7 vs 10/year) and were more likely to use emergency care. Overall, 16% of their care was emergency vs 9% for the least deprived (Table 2).

Finally, people added *ad hoc* locally to the register (who were not initially identified as needing to shield by the Scottish Government) had higher healthcare use pre-pandemic than those who were identified in the national record search. Those identified locally were more likely to have had out-patient attendance (91% vs 79%), scheduled in-patient admissions (32% vs 24%) and emergency in-patient admission (26% vs 23%).

We also analysed the chronic disease burden of the shielding register population during the 5 years pre-pandemic (Table 2). 38% of the shielding patients had more than one chronic disease diagnosed (were multimorbid), with a mean morbidity burden score of 5.6 (Table 2). Multimorbidity and morbidity burden increased with increasing age but not deprivation levels or rurality (Table 2).

### Changes in healthcare use over time in the shielding register population

During the period of 26^th^ March through 18^th^ June 2020 when strict self-isolation was recommended for people on the shielding register, the rates of healthcare use decreased for all types of care: out-patient, both scheduled and emergency in-patient admissions, and A&E (Figure 3).

The total out-patient attendance rate dropped substantially in the lead up to the shielding period, with the lowest rate at the start of the shielding period. At the same time, the proportion of out-patient visits conducted virtually increased substantially, with half of all visits taking place virtually during shielding (Figure 3A). The decrease in out-patient care use varied widely across clinic types (Figure 4) – with cancer care clinics seeing smaller decreases than the other most-attended clinics for shielding people (Figure 4).

**Figure 4.**
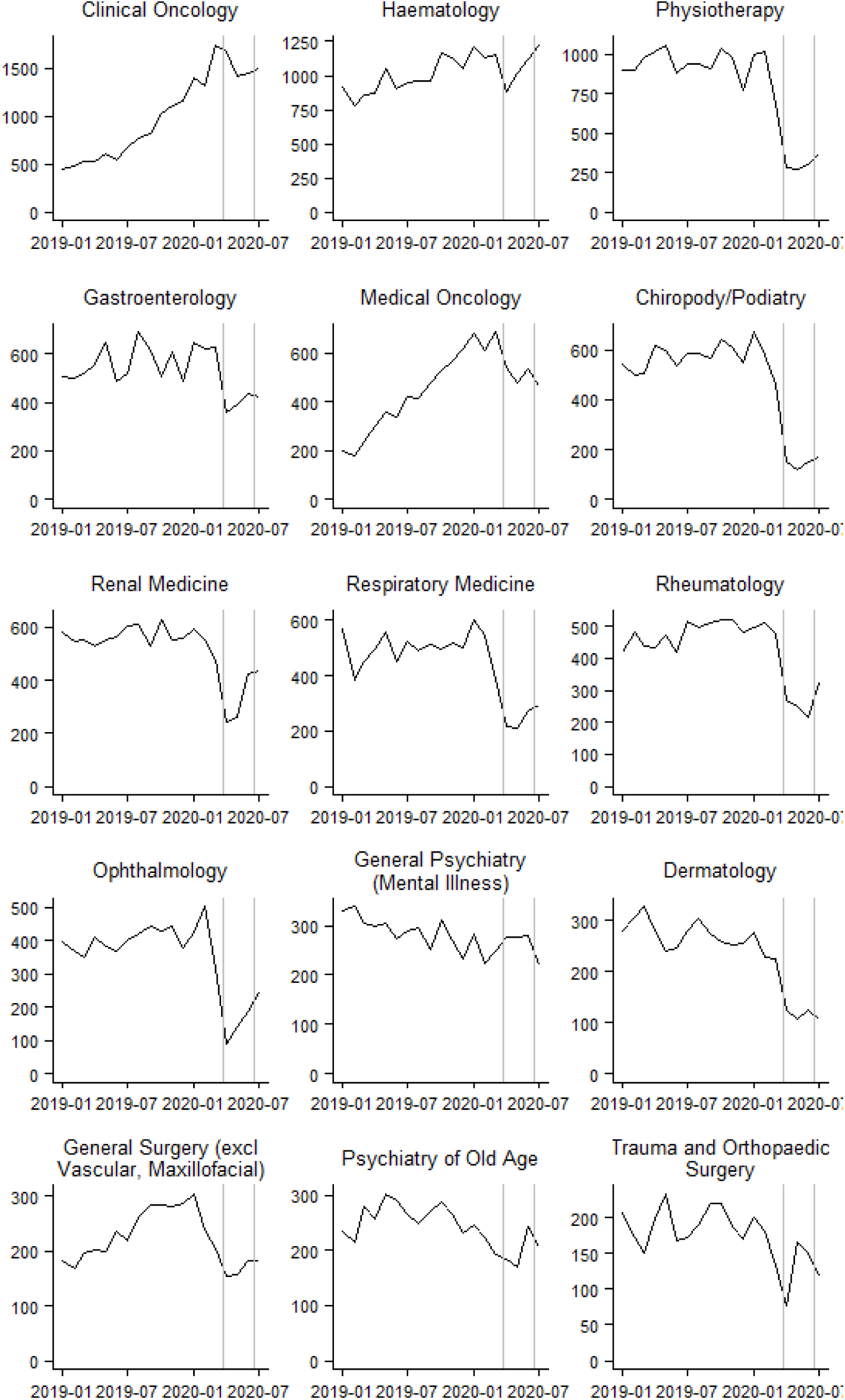
Monthly out-patient care use by clinic type for the shielding register population. Attendances per month for the shielding population at the 16 out-patient clinics with the highest attendances. Gray lines show the shielding period 26^th^ March to 18^th^ June 2020. Clinic names as given in Scottish Morbidity Record 00. *Note the Y axis scale varies per clinic.*

In the two years pre-pandemic, the shielding population consistently had more scheduled than emergency in-patient hospital admissions (Figure 3B). During shielding, this relationship reversed, with scheduled in-patient admissions decrease more than emergency visits (Figure 3B). Emergency department attendances also dropped substantially in the lead up to the start of the shielding period, with their lowest point at the very beginning of the period (Figure 3C).

### Comparison of healthcare use between the shielding and total non-shielding populations

We compared the change in healthcare use between the population on the shielding register (16,092) to the rest of the population in the NHS Grampian region (569,608) before and during the lockdown. Per capita, the shielding population had much higher care use for all types of care (Figure 5). Both shielding and non-shielding populations saw substantial decreases in all types of care use in the period leading up to and during lockdown (Figure 5). The steepest decline was seen in scheduled in-patient care among the non-shielding population (Figure 5d).

**Figure 5.**
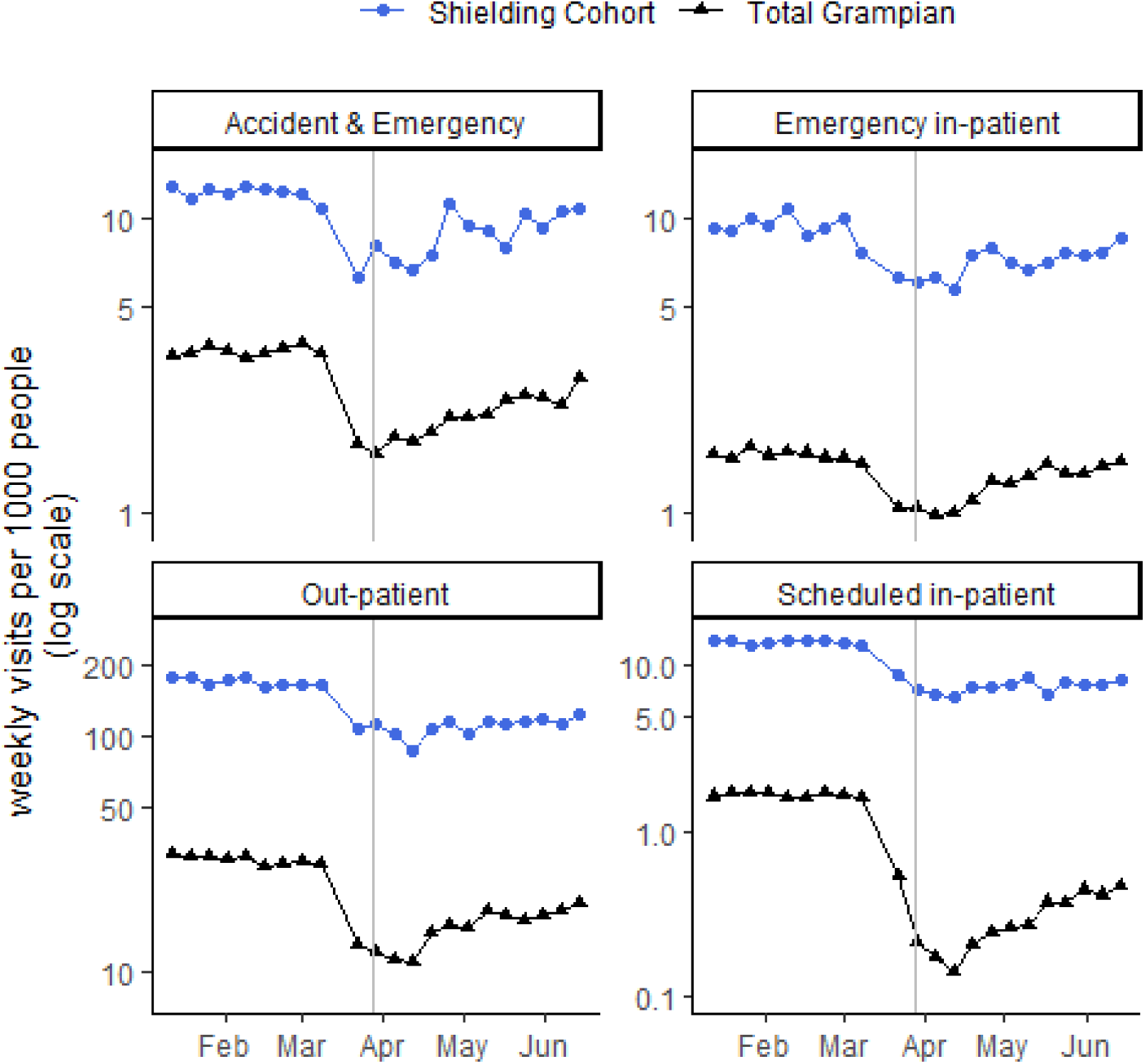
Healthcare use in 2020 by the shielding population compared to the total non-shielding NHS Grampian region population. Weekly attendances and admissions per 1000 people for out-patient, scheduled in-patient, emergency in-patient, and accident and emergency care. Gray line shows the start of the shielding period on 26^th^ March 2020. *Note the Y axis is log scale and varies for each care type*.

We modelled healthcare use by the shielding populations and non-shielding populations before and during lockdown. For the shielding population, scheduled care fell more than emergency care while shielding – the largest decline was seen in scheduled in-patient care, which fell to 54% of its pre-shielding level, out-patient care fell to 65% of its pre-shielding level, emergency in-patient care fell to 75%, and A&E fell to 71% (Table 3).

**Table 3.** Population-level changes in healthcare use for lockdown versus pre-lockdown time periods (expressed as rate ratio) for people shielding and for the general non-shielding population

|  | Shielding population<br>change in healthcare use<br>during lockdown |  | All Grampian non-shielding<br>population change in<br>healthcare use during<br>lockdown |  | Rate ratio for change in<br>healthcare use during<br>lockdown for shielding vs<br>non-shielding |  |
| --- | --- | --- | --- | --- | --- | --- |
|  | RR | CI | RR | CI | RR | CI |
| **Out-patient** | 0.65 | (0.60-0.71) | 0.51 | (0.47-0.56) | 1.27 | (1.13-1.43) |
| **Scheduled in-patient** | 0.54 | (0.45-0.66) | 0.19 | (0.16-0.22) | 2.91 | (2.23-3.78) |
| **Emergency in-patient** | 0.75 | (0.68-0.83) | 0.79 | (0.73-0.84) | 0.96 | (0.85-1.08) |
| **Accident & Emergency** | 0.71 | (0.65-0.78) | 0.59 | (0.55-0.64) | 1.20 | (1.07-1.35) |

Notes: RR > 1 for Shielding vs Grampian non-shielding populations indicates
a more substantial reduction in healthcare for the Grampian population than for those shielding.

However, care levels for the shielding population were better maintained (i.e., reduction in healthcare use was less) compared to the rest of Grampian for all healthcare types (Table 3). Notably, for the non-shielding population scheduled in-patient care fell to 19% of its pre-lockdown level (compared with 54% of pre-lockdown levels for the shielding population) (Table 3). Thus, the shielding population had higher use of scheduled in-patient by three-fold (RR = 2.9, 95% CI = 2.2 – 3.8), out-patient by 30% (RR = 1.3, 95% CI = 1.1-1.4) and A&E care by 20% (RR = 1.2, 95% CI = 1.1 – 1.4) compared with the non-shielding population during the lockdown, with no evidence of difference in the emergency in-patient care (RR = 0.96; 95% CI = 0.9-1.1) (Table 3).

### Description of the shielding population who had healthcare visits during the shielding period

To better understand who on the shielding register used care during the shielding period we described their demographic and clinical characteristics and modelled care use across the sub-populations (Table 4).

**Table 4.** Characteristics of shielding population with healthcare use during the shielding period. Healthcare use between 1^st^ April and 31^st^ July 2020 for the shielding population total, as well as stratified by out-patient, in-patient, A&E care, routine and emergency care users. *Scheduled Care* includes all scheduled care in both out-patient and in-patient settings and *Emergency Care* includes all unscheduled care in both in-patient and A&E settings. Morbidity burden is weighted total chronic disease diagnoses from in-patient admissions in the 5 years from 1^st^ March 2015.

|  | N | Women | Age (med) | People <20 years old | Deprived Home Area | Remote or Rural | Identified Nationally | % Multi-morbid | Morbid Burden (med) |
| --- | --- | --- | --- | --- | --- | --- | --- | --- | --- |
| **TOTAL SHIELDING** | 16092 | 8632 (54.0%) | 65 | 496 (3.0%) | 1230 (8.0%) | 5494 (34.0%) | 9354 (58.0%) | 38% | 5.6 |
| **Any care while shielding** |  |  |  |  |  |  |  |  |  |
| **Care** | 8699 | 4649 (53.0%) | 65 | 387 (4.0%) | 634 (7.0%) | 2917 (34.0%) | 4238 (49.0%) | 46% | 6.5 |
| **No Care** | 7393 | 3983 (54.0%) | 65 | 109 (1.0%) | 596 (8.0%) | 2577 (35.0%) | 5116 (69.0%) | 29% | 4.3 |
| **Out-patient visit** |  |  |  |  |  |  |  |  |  |
| **Visit** | 8179 | 4384 (54.0%) | 64 | 368 (4.0%) | 587 (7.0%) | 2744 (34.0%) | 3871 (47.0%) | 45% | 6.5 |
| **No Visit** | 7913 | 4248 (54.0%) | 65 | 128 (2.0%) | 643 (8.0%) | 2750 (35.0%) | 5483 (69.0%) | 31% | 4.5 |
| **Out-patient visit** |  |  |  |  |  |  |  |  |  |
| **In person** | 1755 | 955 (54.0%) | 64 | 60 (3.0%) | 149 (8.0%) | 558 (32.0%) | 1008 (57.0%) | 46% | 5.4 |
| **Virtual** | 6424 | 3429 (53.0%) | 64 | 308 (5.0%) | 438 (7.0%) | 2186 (34.0%) | 2863 (45.0%) | 45% | 6.8 |
| **Out-patient visit** |  |  |  |  |  |  |  |  |  |
| **New visit** | 2440 | 1349 (55.0%) | 64 | 188 (8.0%) | 195 (8.0%) | 756 (31.0%) | 1156 (47.0%) | 46% | 6.4 |
| **Return** | 6743 | 3552 (53.0%) | 64 | 314 (5.0%) | 459 (7.0%) | 2298 (34.0%) | 2987 (44.0%) | 45% | 6.7 |
| **Other** | 2587 | 1387 (54.0%) | 64 | 69 (3.0%) | 165 (6.0%) | 789 (30.0%) | 1040 (40.0%) | 46% | 7.3 |
| **In-patient visit** |  |  |  |  |  |  |  |  |  |
| **Visit** | 2326 | 1200 (52.0%) | 67 | 133 (6.0%) | 186 (8.0%) | 776 (33.0%) | 1093 (47.0%) | 55% | 8 |
| **No visit** | 13766 | 7432 (54.0%) | 65 | 363 (3.0%) | 1044 (8.0%) | 4718 (34.0%) | 8261 (60.0%) | 36% | 5.1 |
| **In-patient visit** |  |  |  |  |  |  |  |  |  |
| **Emergency** | 1755 | 897 (51.0%) | 68 | 85 (5.0%) | 153 (9.0%) | 535 (30.0%) | 879 (50.0%) | 58% | 8.4 |
| **Scheduled** | 811 | 412 (51.0%) | 63 | 86 (11.0%) | 50 (6.0%) | 323 (40.0%) | 294 (36.0%) | 47% | 7.1 |
| **A&E visit** |  |  |  |  |  |  |  |  |  |
| **Visit** | 1805 | 952 (53.0%) | 69 | 47 (3.0%) | 166 (9.0%) | 552 (31.0%) | 978 (54.0%) | 58% | 7.9 |
| **No Visit** | 14287 | 7680 (54.0%) | 64 | 449 (3.0%) | 1064 (7.0%) | 4942 (35.0%) | 8376 (59.0%) | 36% | 5.3 |

Half the shielding register population (54%) had at least one healthcare visit of any type during the shielding period (Table 4). Compared to those with no healthcare during the shielding period, the group with healthcare visits were more multimorbid (46% vs 29%), younger (4% vs 1% age < 20) and identified *ad hoc* as clinically extremely vulnerable rather than by the national search (51% vs 31%) (Table 4).

Outpatient care was primarily by virtual appointments. 51% of people had an out-patient attendance, but only 11% had an *in-person* out-patient visit, with the rest using phone or video (Table 4). Substantially fewer people had in-patient care: only 5% had a scheduled in-patient admission (Table 4). 11% of people had an emergency in-patient admission and 11% had an A&E attendance during the shielding period (Table 4).

### Association of age, sex, home-area deprivation, and reason for shielding with change in healthcare use

We modelled total healthcare visits for the shielding register population by sex, home-area deprivation, age and reason for shielding for pre-lockdown and during-lockdown phases.

Prior to lockdown, we did not find any evidence that that the healthcare use differed between men and women (Table 5). People from deprived areas had greater emergency care compared to those from non-deprived areas (emergency in-patient care 32% higher and A&E use 52% higher) (Table 5). People shielding due to cancer used substantially greater care than any other shielding category (compared to respiratory patients: 3.2-fold higher out-patient, 5.4-fold higher scheduled in-patient, 2.3-fold higher emergency in-patient and 1.6-fold higher A&E) (Table 5).

**Table 5.** Differences in healthcare use across sub-populations of the shielding register in the period before shielding. RR = ratio of mean healthcare visits from 1 January to 14 March 2020 compared to the reference level as indicated in parentheses, CI = 95% confidence interval.

| Population subset (vs reference) | Out-patient |  | In-patient scheduled |  | In-patient emergency |  | A&E |  |
| --- | --- | --- | --- | --- | --- | --- | --- | --- |
|  | RR | CI | RR | CI | RR | CI | RR | CI |
| women (vs men) | 1.12 | (0.99-1.25) | 0.85 | (0.68-1.05) | 0.97 | (0.81-1.14) | 1.01 | (0.88-1.16) |
| deprived (vs not) | 1.07 | (0.95-1.20) | 1.03 | (0.80-1.33) | 1.32 | (1.05-1.66) | 1.53 | (1.26-1.85) |
| <50 years old (vs 80+) | 1.09 | (0.91-1.31) | 1.12 | (0.79-1.58) | 1.07 | (0.82-1.40) | 0.81 | (0.64-1.02) |
| 50-59 | 0.93 | (0.77-1.12) | 0.80 | (0.55-1.15) | 0.64 | (0.47-0.86) | 0.73 | (0.57-0.93) |
| 60-69 | 1.02 | (0.85-1.23) | 1.14 | (0.80-1.62) | 0.71 | (0.54-0.94) | 0.82 | (0.66-1.03) |
| 70-79 | 1.04 | (0.87-1.26) | 1.19 | (0.84-1.69) | 0.85 | (0.65-1.11) | 0.78 | (0.63-0.98) |
| cancer (vs respiratory) | 3.22 | (2.68-3.87) | 5.38 | (3.93-7.35) | 2.31 | (1.85-2.90) | 1.55 | (1.29-1.87) |
| immunosuppressants | 1.44 | (1.19-1.74) | 1.65 | (1.17-2.32) | 0.55 | (0.42-0.74) | 0.55 | (0.44-0.69) |
| other | 1.43 | (1.18-1.73) | 2.02 | (1.43-2.85) | 1.16 | (0.88-1.51) | 0.92 | (0.73-1.16) |
| rare disease | 1.26 | (1.03-1.54) | 0.94 | (0.61-1.45) | 0.74 | (0.52-1.07) | 0.64 | (0.46-0.87) |
| transplant | 1.99 | (1.61-2.45) | 2.36 | (1.55-3.60) | 1.52 | (1.06-2.18) | 1.07 | (0.77-1.49) |

Compared to the pre-shielding period, all subgroups of the shielding population had a substantial reduction of healthcare use for all types of care (ratio of mean visits < 1 in Table 6). Scheduled care was affected more than emergency care across all subsets of the shielding population (ratio of mean visits smaller for scheduled vs emergency care types in Table 6).

**Table 6.**
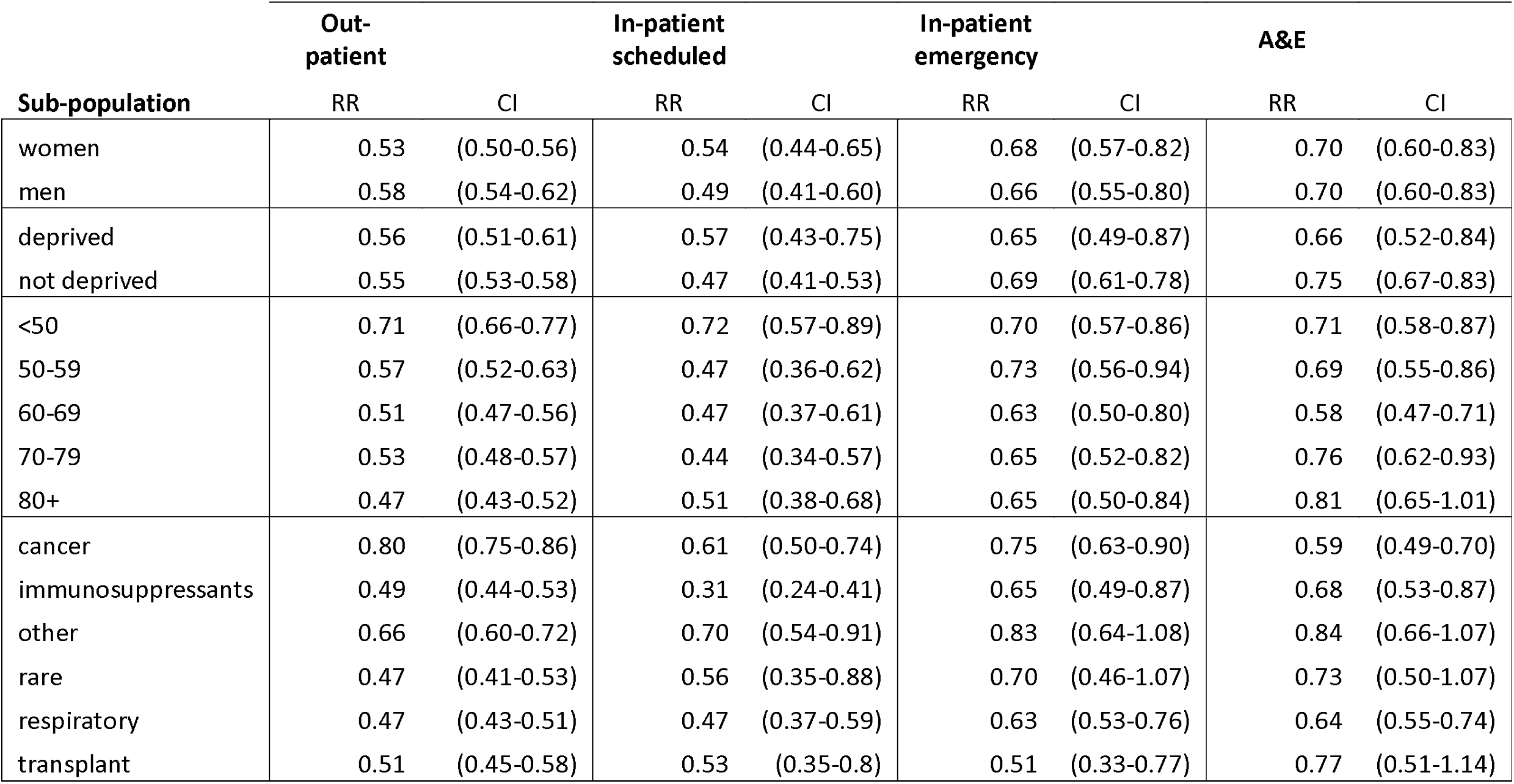
Differences in healthcare use across sub-populations of the shielding register before and during shielding. RR = ratio of mean healthcare visits from 22 March to 18 June 2020 compared to 1 January to 14 March 2020, CI = 95% confidence interval

There was no evidence of a difference in care between men and women, or between people who lived in deprived home-areas and those who did not (Table 6). All had scheduled care rates of about half of pre-lockdown levels and emergency care of about two-thirds of pre-lockdown levels (Table 6).

There were significant differences in care use change across reasons for shielding. People with cancer had better maintained out-patient activity during lockdown – 80% of pre-lockdown levels, compared to ∼50% seen for people shielding for other reasons (Table 6). There was also preservation of all scheduled care for younger people – both out-patient and scheduled in-patient care rates were higher during lockdown than those of all other age groups, at over 70% of pre-lockdown levels while it was ca. 50% for older age groups (Table 6).

Comparison of predicted mean visits between pre-shielding and shielding by age group and reason for shielding are presented graphically (Figure 6 and Supplementary File).

**Figure 6.**
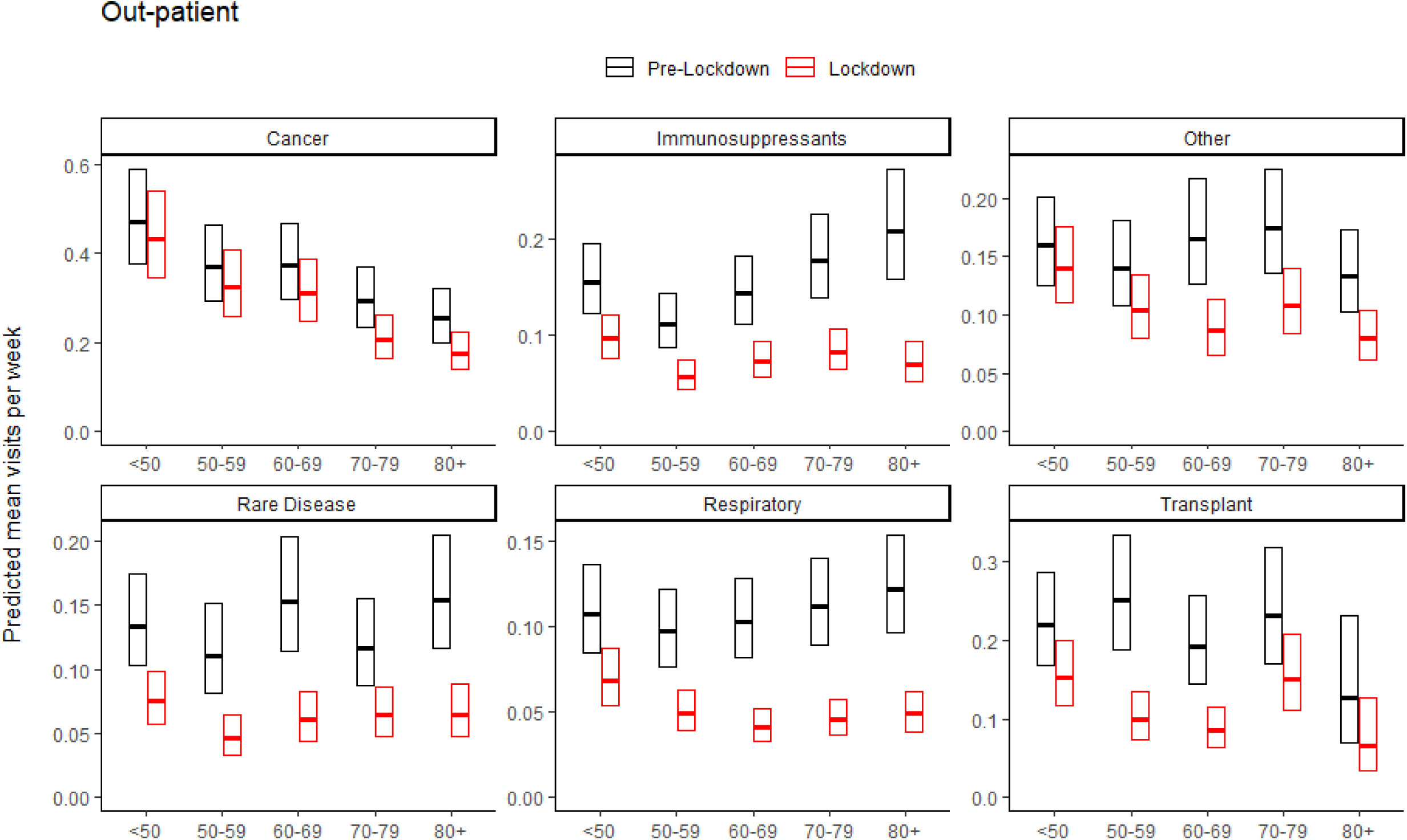
Comparison of mean out-patient visits pre-lockdown and during lockdown by age and reason for shielding. Pre-lockdown period is 1 January to 14 March 2020 in black and the lockdown period is 22 March to 18 June 2020 in red. *Note the Y axis scale varies for each care type.*

While shielding, people of all ages had fewer healthcare visits for all types of care and increased reliance on emergency care regardless underlying reason for shielding (Figure 6 and Supplementary File). Across all reasons for shielding, the youngest people saw the best maintenance of out-patient care rates (Figure 6) and across all ages, people shielding due to cancer saw the best maintained out-patient care (Figure 6).

## DISCUSSION

The objective of the shielding programme was to: 1) identify those most clinically vulnerable to COVID, 2) offer support during self-isolation, 3) protect them from emergency healthcare contact that could lead to infection while 4) maintaining essential care for pre-existing illnesses. No population study has comprehensively evaluated how the clinically extremely vulnerable were identified and how their healthcare changed while shielding. The research presented here shows that many people considered to be clinically extremely vulnerable were not captured by electronic health record screening, despite their high healthcare use. We also found that for shielding people, scheduled care was maintained to a greater extent than the general population, but that there was significant variation in care maintenance within the shielding population.

We found that Grampian had a smaller proportion of its population advised to shield (2.7%) than the Scottish average (3.3%), which was smaller than other UK countries (4.0% in England and 4.2% in Wales)^23^. Notably, 42% of Grampian’s shielding population was not identified as clinically extremely vulnerable in the Scottish health record screen and instead had to be added *ad hoc* by local clinicians, in contrast with 22% added *ad hoc* in Wales^23^. A contributing factor in both of these differences may be the lack of integration of primary care records in the automated screen for the clinically extremely vulnerable in Scotland compared to England and Wales. The *ad hoc* additions in Grampian were a younger group of people with similar morbidity but higher healthcare use, who live in areas of less deprivation. The large number who had to be added and the socioeconomic differences found in those who were added *ad hoc*, indicate that the lack of integration of data when defining the most vulnerable may lead to less equitable care prioritisation. If identification of the vulnerable was suboptimal, then access to shielding support and risk mitigation will have been suboptimal.

Those on the shielding register had exceptionally high (and rising) healthcare use prior to the pandemic due to their serious underlying health conditions. As with the general population, the shielding population’s overall healthcare use declined rapidly leading up to the first lockdown. However, encouragingly, while emergency care dropped to a similar extent for shielding and non-shielding people, scheduled care was better maintained for those on the shielding register. Overall, this implies that for those with particularly high care needs, the continuity of scheduled care was somewhat protected, although there was substantial variability in different subsets of the shielding population. Those who were young and those with cancer had better maintained scheduled care, whereas those who were older or shielding for other reasons experienced larger reductions. We also found that before the pandemic there was a socioeconomic gradient in access to scheduled care which did not change during the lockdown, despite evidence that people living in deprived areas being at excess risk of poor outcomes^26,27^. This work demonstrates the feasibility of maintaining continuity of care for priority at-risk groups and suggests prioritisation of care should include sociodemographic information.

This analysis complements two recent studies from the UK showing that people who shield had high emergency care use, an increased risk of nosocomial coronavirus infection, and poorer outcomes than the general population^28,29^. These studies’ results underline the need to minimise emergency care where possible without compromising continuity of care for pre-existing conditions. Our analysis shows that this was feasible, particularly where continuity was prioritised (such as in cancer), but further evaluation is necessary to understand if other sub-populations should also be prioritised given evidence of poorer outcomes among people from ethnic minorities and those living in deprived areas^26,27^.

Strengths of this study include the whole population design, with capture of all people on the shielding register during the lockdown period, and the use of pre-lockdown comparisons across a wide variety of in-patient and out-patient care. It also includes a comparison with the general population to contextualise the changes seen in the shielding register population. Limitations include that the population covers a region that was somewhat less affected by coronavirus (COVID death rates at the end of the shielding period of 31^st^ July 2020: 26 per 100,000 in Grampian compared to 47/100,000 in Scotland, 54/100,000 in Lothian and 63/100,000 in Greater Glasgow and Clyde^30^). An appropriate next step would be to scale these analyses across wider areas of Scotland and the UK.

Rapid and reliable identification of the clinically vulnerable will continue to be important, and this analysis suggests two ways identification could be improved. First, sharing primary care records. In Scotland, primary care records are not shared nationally, limiting who could be identified as clinically extremely vulnerable to COVID and supported. Second, improving person-level sociodemographic data collection, including ethnicity. COVID hospital admissions and deaths have made it clear that sociodemographic characteristics affect clinical vulnerability to COVID. But in Scotland, ethnicity data are not well recorded, and the sociodemographic data available is for small areas rather than individuals. Neither ethnicity no socioeconomic data were not used to identify those who should shield. Collecting these data during healthcare visits and sharing them nationally could help improve care of the people who are clinically vulnerable.

Healthcare changed dramatically for the clinically extremely vulnerable population during the pandemic. The increased reliance on emergency care while isolating indicates that continuity of care for existing conditions was not optimal. However, compared to the general population, there was success in maintaining scheduled care, particularly in young people and those with cancer. We suggest that integrating demographic and primary care data would improve identification of the clinically vulnerable and could aid prioritising their care.

## Supporting information

Analysis R code

Authors' Credit Taxonomy

## Data Availability

All analysis was carried out in the Grampian Data Safe Haven (project ID: DaSH412) on pseudonomysed individual-level data. Per UK General Data Protection Regulation, only aggregate data can be released from the Grampian Data Safe Haven for publication, but all individual-level data has been archived and can be accessed by application to the Grampian Data Safe Haven.

## ACKNOWLEDGMENTS

This work was funded by the Scottish Government’s Chief Scientist Office via the Rapid Research in COVID-19 Programme and by⍰the Health Foundation via the Networked Data Lab Programme. The Health Foundation is an independent charity⍰committed to bringing about better health and healthcare for people⍰in the UK. We thank the team at the Grampian Data Safe Haven facility (DaSH) within the Aberdeen Centre for Health Data Science and the associated financial support of DaSH by the University of Aberdeen and NHS Research Scotland through NHS Grampian investment. This work uses data provided by patients and collected by the NHS as part of their care and support.

