## Supplementary material for "The clinically extremely vulnerable to COVID: Identification and changes in healthcare while self-isolating (shielding) during the coronavirus pandemic": Analysis R code

Analysis Code

\# Libraries

```{r, message = F}

library(tidyverse)

library(lubridate)

library(comorbidity)

library(cowplot)

library(janitor)

library(vroom)

library(here)

options(scipen = 999)

theme_set(theme_cowplot())

```

\# Shielding list

```{r message = F}

#load and tidy shielding list

#this file has individuals listed one time *for each reason shielding*

shielding_original <-

vroom(here("original_data", 'Dash416_Shield20201012_Release.csv'),

delim = "¬") %>%

clean_names() %>%

rename(id = dash416_release_uid) %>%

mutate(date_addition =

as_date(ymd_hms(earliest_addition_this_chi)),

date_removal =

as_date(ymd_hms(most_recent_removal_date)),

date_death = as_date(ymd(trak_person_deceased_date))) %>%

select(id,

group,

origin_for_this_group,

date_addition,

date_removal,

removal_description,

date_death) %>%

distinct()

#add translation of shielding group codes

shielding_original <-

vroom(here("original_data", "shielding_codes_short.csv"),

delim = ",") %>%

left_join(shielding_original, ., by = c("group" = "shielding_group"))

```

\# Population exclusions

```{r}

#anyone who died before 26 March (first day of shielding)

#false positives from a lung cancer screen

#moved from Scotland

shielding_long <-

shielding_original %>%

filter(date_death > ymd("2020-03-26") |

is.na(date_death)) %>%

filter(

!removal_description %in%

c("FalsePosLungCancer", "False Positive", "Moved out of Scotland"))

#vector of ids of people in final cohort for paper

unique_ids <- unique(shielding_long$id)

n_shielders = length(unique_ids)

```

#Unique shielding list

```{r}

#concatenate all individuals' group membership

shielding_groups <-

shielding_long %>%

group_by(id) %>%

arrange(group) %>%

summarise(

shielding_groups = paste(group, collapse = ", "),

shielding_reasons = paste(shielding_group_description, collapse = ", "))

#add variable of source - if added at national record scan for any reason then national, else local

shielding_source <-

shielding_long %>%

group_by(id) %>%

summarise(

central = sum(origin_for_this_group == "central"),

local = sum(origin_for_this_group == "local")) %>%

mutate(source = ifelse(central > 0, "National", "Local")) %>%

select(id, source)

#unique patient characteristics

shielding_people <-

shielding_long %>%

group_by(id) %>%

slice_head() %>%

select(

id,

date_addition,

date_removal,

date_death

) %>%

left_join(., shielding_source, by = "id") %>%

left_join(., shielding_groups, by = "id")

#add single shielding category for people who have multiple

#use hierarchy of cancer, transplant, respiratory, rare, immune, other (includes pregnant)

shielding_people <-

shielding_people %>%

mutate(

dominant_reason =

case_when(

str_detect(shielding_reasons, "Cancer") ~ "Cancer",

str_detect(shielding_reasons, "Transplant") ~ "Transplant",

str_detect(shielding_reasons, "Respiratory") ~ "Respiratory",

str_detect(shielding_reasons, "Rare") ~ "Rare disease",

str_detect(shielding_reasons, "Immunosuppressants") ~ "Immunosuppressants",

TRUE ~ "Other"

)

)

rm(shielding_original, shielding_groups, shielding_source)

```

\# Demographic data

```{r}

#load demographic data

demog <-

vroom(here("original_data", 'Dash416_Shield20201012_Demographic_Release.csv'),

delim = "¬") %>%

rename(id = Dash416_Release_UID) %>%

clean_names() %>%

select(id, calc_sex, calc_dob)

#load small area statistics about home

vulnerability <-

vroom(here("original_data", 'Dash416_Shield20201012_vulnerability_Release.csv'),

delim = "¬") %>%

clean_names() %>%

rename(id = dash416_release_uid,

ur_name = u_rname,

ur_class = u_rclass,

simd_decile = simd2020v2_decile,

simd_vigntile = simd2020v2_vigintile,

simd_rank = simd2020v2_rank) %>%

#mutate(simd_vigntile = as_factor(simd_vigntile)) %>%

select(id, simd_decile, simd_vigntile, simd_rank, ur_class, ur_name)

#keep only people in cohort

demog <-

demog %>%

filter(id %in% unique_ids)

#create variables for sex and age band

#dob is only given as 1st of month of year born (disclosure control)

#age is calculated as of 1st March 2020

demog <- demog %>%

mutate(

sex = if_else(calc_sex == 1, "F", "M"),

dob = ymd(calc_dob),

age = as.period(interval(dob, ymd("2020-03-01")), units = "years")$year,

age_band =

case_when(

age %in% c(0:19) ~ "0-19",

age %in% c(20:29) ~ "20-29",

age %in% c(30:39) ~ "30-39",

age %in% c(40:49) ~ "40-49",

age %in% c(50:59) ~ "50-59",

age %in% c(60:69) ~ "60-69",

age %in% c(70:79) ~ "70-79",

age %in% c(80:89) ~ "80-89",

age >=90 ~ "90+")) %>%

select(-calc_sex,-calc_dob)

#set age band as ordered factor

demog$age_band <- factor(demog$age_band, levels = c("0-19", "20-29", "30-39", "40-49", "50-59", "60-69", "70-79", "80-89", "90+"))

#add details of home area to demographics

demog <-

demog %>%

left_join(., vulnerability, by = "id")

#add deaths to demog

demog <-

shielding_people %>%

left_join(demog, ., by = "id") %>%

mutate(dead = ifelse(is.na(date_death), 0, 1)) %>%

relocate(date_death, .after = last_col())

#add some demographics values to long shielding list

shielding_long <-

demog %>%

select(id, age, age_band, sex, simd_decile, ur_name) %>%

left_join(shielding_long, ., by = "id")

rm(vulnerability, shielding_people)

#end result is two files: one (demog) with one case per person, and one (shielding_long) with one case per reason shielding

```

\# A&E data

```{r}

ae <-

vroom(here(

"original_data",

'Dash416_HI_ED_Attendances_Release_v2_1.csv'

),

delim = "¬") %>%

clean_names() %>%

rename(id = dash416_release_uid,

date = arrival_date)

#filter for cohort

ae <-

filter(ae,

id %in% unique_ids)

#add demog data to A&E

ae <-

ae %>%

left_join(., demog, by = "id")

```

#A&E 1 yr

```{r}

demog <-

ae %>%

filter(date >= ymd("2019-03-01"),

date < ymd("2020-03-01")) %>%

group_by(id) %>%

summarise(n_ae_1yr = n(),

n_ae_major_1yr = sum(major_minor == "Major"),

.groups = "drop") %>%

left_join(demog, ., by = "id")

```

\# Outpatient data

```{r}

#load and tidy Trak outpatient data

op <-

vroom(here("original_data", 'Dash416_HI_OP_Admissions_Release_v2_1.csv'), delim = "¬") %>%

clean_names() %>%

rename(id = dash416_release_uid,

date = appointment_date,

virtual = virtual_non_virtual_appts,

specialty = specialty_local_desc,

specialty_national = specialty_national_desc)

#exclusions for cohort and attended

op <-

op %>%

filter(id %in% unique_ids) %>%

filter(attendance_status_desc != "DNA") %>%

select(-attendance_status_desc)

#add demog data to op

op <-

op %>%

left_join(., demog, by = "id")

```

\# Out-patient 1 & 5 year

```{r}

demog <-

op %>%

filter(date >= ymd("2019-03-01"),

date < ymd("2020-03-01")) %>%

group_by(id) %>%

summarise(n_op_1yr = n(),

.groups = "drop") %>%

left_join(demog, ., by = "id")

demog <-

op %>%

filter(date >= ymd("2015-03-01"),

date < ymd("2020-03-01")) %>%

group_by(id) %>%

summarise(different_clinics_5yr = length(unique(specialty_national)),

.groups = "drop") %>%

left_join(demog, ., by = "id")

```

\# SMR inpatient data

```{r}

#load data, rename variables, format dates, add episode ID and date month, select only variables needed

smr01_long_original <-

vroom(here("original_data", 'Dash416_Shield20201012_SMR01_Release.csv'),

delim = "¬") %>%

clean_names() %>%

rename(id = dash416_release_uid,

main_condition = main_cod,

other_condition1 = oc1,

other_condition2 = oc2,

other_condition3 = oc3,

other_condition4 = oc4,

other_condition5 = oc5) %>%

mutate(

date_episode_start = as_date(ymd(date_episode_start)),

date_episode_end = as_date(ymd(date_episode_end)),

date_admission = as_date(ymd(adm_date)),

date_discharge = as_date(ymd(date_discharge)),

episode_id = paste(id, date_episode_start, sep = "_"))

#select date range and those in population

smr01_long <-

filter(smr01_long_original,

date_episode_start >= ymd("2018-01-01"),

id %in% unique_ids)

```

\# SMR01 single episode

```{r}

smr01_flags<-

smr01_long %>%

group_by(episode_id) %>%

summarise(emergency = sum(emerg),

itu = sum(itu),

shdu = sum(shdu),

ccu = sum(ccu),

from_care = sum(loc_admission),

to_care = sum(loc_discharge),

.groups = "drop")

smr01_los <-

smr01_long %>%

group_by(episode_id) %>%

slice_head() %>%

ungroup() %>%

select(id, episode_id, date_episode_start, date_episode_end) %>%

mutate(episode_los =

difftime(date_episode_end, date_episode_start, units = "days"))

smr01 <-

left_join(smr01_los, smr01_flags, by = "episode_id")

#add demog data to hospital admissions

smr01 <-

smr01 %>%

mutate(date = date_episode_start) %>%

left_join(., demog, by = "id")

rm(smr01_flags, smr01_los)

```

\# SMR01 1 yr

```{r}

demog <-

smr01 %>%

filter(date >= ymd("2019-03-01"),

date < ymd("2020-03-01")) %>%

group_by(id) %>%

summarise(n_smr01_1yr = n(),

n_emerg_1yr = sum(emergency == 1),

n_routine_1yr = n_smr01_1yr - n_emerg_1yr,

los_smr01_1yr = sum(episode_los),

.groups = "drop") %>%

left_join(demog, ., by = "id")

```

\# Tidy 1 year care

```{r}

#change na to 0

#sum op + routine ip as "routine" and ae + emergency ip as "emergency"

demog <-

demog %>%

mutate(

across(n_ae_1yr:los_smr01_1yr, ~ replace_na(., 0)),

all_routine_1yr = n_op_1yr + n_routine_1yr,

all_emergency_1yr = n_ae_1yr + n_emerg_1yr,

prop_care_emergency_1yr =

round(all_emergency_1yr / (all_emergency_1yr + all_routine_1yr), 2))

```

\# Morbidity

```{r}

#make long df that has one diagnosis per row (comorbidity input)

#for 5 years before lockdown

smr01_diagnoses <-

smr01_long_original %>%

filter(date_episode_start >= ymd("2015-03-01"),

date_episode_start < ymd("2020-03-01"),

id %in% unique_ids) %>%

pivot_longer(main_condition:other_condition5, values_drop_na = T) %>%

select(id, episode_id, date_episode_start, name, value)

#use comorbidity package to calculate chronic diseases

person_morbidity <-

comorbidity(

x = smr01_diagnoses,

id = "id",

code = "value",

score = "elixhauser",

icd = "icd10",

assign0 = T

) %>%

mutate(score_band = case_when(score == 0 ~ 0,

score == 1 ~ 1,

score > 1 ~ 2))

#add chronic disease data to demographics

demog <-

demog %>%

left_join(., person_morbidity, by = "id")

rm(person_morbidity, smr01_long_original, smr01_diagnoses)

```

#Grampian data

```{r}

#in-patient

gramp_ip <-

vroom(here("original_data", 'vulnerable_416_All_IP_Admsssions_Aggregate.csv'), delim = "¬") %>%

select(-Visit_Status_Desc) %>%

rename(date = AdmDate,

type = Adm_Type,

total = no_adms) %>%

mutate(type = ifelse(type == "Non-Elective", "emergency", "scheduled"))

#out-patient

gramp_op <- vroom(here("original_data", 'vulnerable_416_All_OP_Attendances_Aggregate.csv'), delim = "¬") %>%

select(-Attendance_Status_Desc) %>%

rename(date = Appointment_Date,

virtual = "Virtual/Non Virtual appts",

total = no_appts) %>%

mutate(virtual = ifelse(virtual == 1,"virtual", "in person"))

#emergency

gramp_ae <- vroom(here("original_data", 'vulnerable_416_All_ED_Admissions_Aggregate.csv'), delim = ",") %>%

select(-Visit_Type_Desc) %>%

select(Arrival_Date, everything()) %>%

rename(date = Arrival_Date,

department = Emergency_Dept_Desc,

arrival = Arrival_Mode,

admitted = "Hospital admission",

total = ED_Atttend)

```

#Time periods

```{r}

#function to fix dates and calculate time periods

fix_dates <-

function(input_file) {

mutate(input_file,

date = as_date(date),

date_week = floor_date(date, unit = "week"),

date_month = floor_date(date, unit = "month"),

date_season = floor_date(date, unit = "season"),

date_year = floor_date(date, unit = "year"))

}

op <- fix_dates(op)

smr01 <- fix_dates(smr01)

ae <- fix_dates(ae)

gramp_op <- fix_dates(gramp_op)

gramp_ip <- fix_dates(gramp_ip)

gramp_ae <- fix_dates(gramp_ae)

```

#Grampian summary

```{r}

gramp_op %>%

filter(date >= ymd("2019-03-01"),

date < ymd("2020-03-01")) %>%

summarise(n_gramp_op_1yr = sum(total))

gramp_ip %>%

filter(date >= ymd("2019-03-01"),

date < ymd("2020-03-01")) %>%

group_by(type) %>%

summarise(n_gramp_ip_1yr = sum(total), .groups = "drop")

gramp_ae %>%

filter(date >= ymd("2019-03-01"),

date < ymd("2020-03-01")) %>%

summarise(n_gramp_ae_1yr = sum(total))

```

#Usage summaries

```{r}

smr01_admissions <-

smr01 %>%

group_by(date_month) %>%

summarise(total = n(),

emergency = sum(emergency),

routine = total - emergency,

.groups = "drop") %>%

mutate(across(total:routine,

.fns = ~./n_shielders * 100,

.names = "{col}_per_100")) %>%

mutate(across(contains("per_100"), round, 1))

op_attendances <-

op %>%

group_by(date_month) %>%

summarise(n_attendances = n(),

n_virtual = sum(virtual),

n_new = sum(referral_type == "New"),

n_return = sum(referral_type == "Return"),

n_adhoc = sum(referral_type == "Adhoc"),

.groups = "drop")

specialties_2019 <-

op %>%

filter(date_year == "2019-01-01") %>%

filter(specialty_national != "Electrocardiography") %>%

count(specialty_national, sort = T) %>%

slice(1:15)

top_15 <- specialties_2019 %>% slice(1:15) %>% .$specialty_national

op_clinics <-

op %>%

filter(specialty_national %in% top_15) %>%

group_by(date_month, specialty_national) %>%

summarise(n = n(),

.groups = "drop")

ae_admissions <-

ae %>%

group_by(date_month) %>%

summarise(total = n(),

major = sum(major_minor == "Major"),

minor = sum(major_minor == "Minor"),

.groups = "drop")

```

#Lockdown usage per person

```{r}

demog <-

smr01 %>%

filter(date >= as_date("2020-04-01")) %>%

filter(date < as_date("2020-08-01")) %>%

group_by(id) %>%

summarise(n_lockdown_admissions = n(),

n_lockdown_admissions_emerg = sum(emergency),

n_lockdown_admissions_routine = sum(emergency == 0),

n_lockdown_admissions_itu = sum(itu),

n_lockdown_admissions_shdu = sum(shdu),

n_lockdown_admissions_ccu = sum(ccu),

.groups = "drop") %>%

left_join(demog, ., by = "id")

demog <-

op %>%

filter(date >= as_date("2020-04-01")) %>%

filter(date < as_date("2020-08-01")) %>%

group_by(id) %>%

summarise(

n_lockdown_op = n(),

n_lockdown_op_virtual = sum(virtual),

n_lockdown_op_new = sum(referral_type == "New", na.rm = T),

n_lockdown_op_return = sum(referral_type == "Return", na.rm = T),

n_lockdown_op_adhoc = sum(referral_type == "Adhoc", na.rm = T),

n_lockdown_op_specialties = n_distinct(specialty),

.groups = "drop") %>%

left_join(demog, ., by = "id")

demog <-

ae %>%

filter(date >= as_date("2020-04-01")) %>%

filter(date < as_date("2020-08-01")) %>%

group_by(id) %>%

summarise(n_lockdown_ae = n(),

n_lockdown_ae_major = sum(major_minor == "Major"),

n_lockdown_ae_hosp = sum(str_detect(departure_destination_desc, "NHS")),

.groups = "drop") %>%

left_join(demog, ., by = "id")

#is this right?

demog <-

demog %>%

mutate(

across(contains("lockdown"), ~ replace_na(., 0)))

```

#Figures

#Source figs

```{r}

source_fig <-

demog %>%

mutate(dominant_reason = fct_rev(fct_infreq(dominant_reason))) %>%

rename(Identified = source) %>%

mutate(Identified = ifelse(Identified == "National", "Nationally", "Grampian")) %>%

ggplot(aes(x = dominant_reason, fill = Identified)) + geom_bar() +

scale_fill_manual(values = c("black", "gray30")) +

coord_flip() + labs(x = "", y = "") + theme(legend.position = c(0.7, 0.2), axis.text = element_text(size = 14))

simd_source <-

demog %>%

filter(!is.na(simd_rank)) %>%

rename(Identified = source) %>%

mutate(Identified = ifelse(Identified == "National", "Nationally", "Locally")) %>%

ggplot(aes(x = simd_rank, fill = Identified)) +

geom_histogram(bins = 50, position = "fill") +

scale_fill_manual(values = c("dark gray", "black")) +

labs(y = "Home Area Deprivation Rank", x = "") +

scale_x_reverse(

breaks = c(275, 2500, 5000, 7000),

labels = c("1\nmost\ndeprived", "2500", "5000", "7000\nleast\n deprived")

) + labs(y = "Proportion identified", x = "Home Area Deprivation Rank") + theme(legend.position = c(0.75, 0.2), legend.key = element_rect(fill = "white"), legend.text = element_text(color = "white"), legend.title = element_text(color = "white"))

age_source_fig <-

demog %>%

filter(age <= 99) %>%

rename(Identified = source) %>%

mutate(Identified = ifelse(Identified == "National", "Nationally", "Locally")) %>%

ggplot(aes(x = age, fill = Identified)) +

geom_histogram(binwidth = 2, position = "fill") +

labs(y = "Proportion identified", x = "Age") +

scale_fill_manual(values = c("dark gray", "black")) +

theme(legend.position = c(0.75, 0.2), legend.key = element_rect(fill = "white"), legend.text = element_text(color = "white"), legend.title = element_text(color = "white"))

```

```{r, fig.height=8}

plot_grid(source_fig, age_source_fig, simd_source,

labels = "AUTO")

```

#Demog figs

```{r}

sex_fig <-

demog %>%

group_by(dominant_reason, sex) %>%

summarise(n = n(),

.groups = "drop") %>%

mutate(dominant_reason = fct_reorder(dominant_reason, n)) %>%

ggplot(aes(x = dominant_reason, y = n, fill = sex)) +

geom_bar(stat = "identity", position= "dodge")+

coord_flip() +

labs(x = "", y = "People") +

theme(legend.position = c(0.8, 0.2), legend.title = element_blank()) +

scale_fill_manual(values = c("gray40", "gray")) +

guides(fill = guide_legend(reverse = T))

age_fig <-

demog %>%

mutate(dominant_reason = fct_reorder(dominant_reason, age)) %>%

ggplot(aes(y = age, x = dominant_reason, fill = sex)) +

geom_boxplot(width = 0.5, size = 1, outlier.shape = NA, coef = 0) +

coord_flip(ylim = c(0, 90)) +

labs(x = "", y = "Age") +

theme(legend.position = c(0.25, 0.9), legend.title = element_blank()) +

scale_fill_manual(values = c("gray", "white")) +

guides(fill = guide_legend(reverse = T))

simd_fig <-

demog %>%

filter(!is.na(simd_decile)) %>%

ggplot(aes(simd_decile, fill = dominant_reason)) +

geom_bar(position = "fill") +

scale_x_reverse(

breaks = c(1, 10),

labels = c("Most\ndeprived", "Least\ndeprived")) +

theme(legend.title = element_blank()) +

labs(y = "Proportion", x = "Home Area Deprivation Deciles")

```

```{r, fig.height=8, fig.width=10}

top_row <- plot_grid(sex_fig, age_fig, labels = c("A", "B"))

plot_grid(top_row, simd_fig, ncol = 1, labels = c("", "C"))

```

#Emerg Routine fig

```{r}

visit_type_reason_fig <-

demog %>%

select(id, dominant_reason, all_emergency_1yr, all_routine_1yr) %>%

rename(Emergency = all_emergency_1yr, Scheduled = all_routine_1yr) %>%

pivot_longer(-c(id, dominant_reason), names_to = "care_type", values_to = "visits") %>%

group_by(dominant_reason, care_type) %>%

summarise(n = sum(visits),

n_people = n(),

mean = mean(visits),

.groups = "drop") %>%

mutate(dominant_reason = fct_reorder(dominant_reason, mean),

care_type = factor(care_type, levels = c("Scheduled", "Emergency"))) %>%

ggplot() +

#geom_point(aes(x = dominant_reason, y = mean), size = 2) +

geom_bar(aes(x = dominant_reason, y = mean), stat = "identity") +

labs(x = "", y = "Pre-pandemic\n annual visits\nper person\n") +

facet_wrap(~care_type, scales = "free") +

theme(legend.position = "none", strip.background = element_rect(fill = "white")) +

theme(axis.text.x = element_text(angle = 35, hjust = 1)) +

expand_limits(y = 0)

visit_type_age_fig <-

demog %>%

select(id, age_band, all_emergency_1yr, all_routine_1yr) %>%

rename(Emergency = all_emergency_1yr, Scheduled = all_routine_1yr) %>%

pivot_longer(-c(id, age_band), names_to = "care_type", values_to = "visits") %>%

group_by(age_band, care_type) %>%

summarise(n = sum(visits),

n_people = n(),

mean = mean(visits),

.groups = "drop") %>%

mutate(care_type = factor(care_type, levels = c("Scheduled", "Emergency"))) %>%

ggplot() +

geom_bar(aes(x = age_band, y = mean), stat = "identity") +

labs(x = "", y = "Pre-pandemic\n annual visits\nper person\n") +

facet_wrap(~care_type, scales = "free") +

theme(legend.position = "none", strip.background = element_rect(fill = "white")) +

theme(axis.text.x = element_text(angle = 45, hjust = 1)) +

expand_limits(y = 0)

```

```{r, fig.height=8, fig.width=8}

plot_grid(visit_type_reason_fig, visit_type_age_fig, ncol = 1, labels = "AUTO", rel_heights = c(1.2, 1))

```

#SMR01 fig

```{r}

#smr01_fig <-

smr01_admissions %>%

filter(date_month <= ymd("2020-07-01")) %>%

pivot_longer(cols = -date_month,

names_to = "admission_type",

values_to = "admissions") %>%

filter(admission_type %in% c("emergency", "routine")) %>%

ggplot(aes(x = date_month, y = admissions, color = admission_type)) +

geom_vline(xintercept = as_date("2020-03-26"), color = "grey") +

geom_vline(xintercept = as_date("2020-06-18"), color = "grey") +

geom_line(aes(linetype = admission_type), size = 1, color = "black") +

ylim(0, 100) +

labs(x = "", y = "In-patient admissions / month") +

scale_linetype_manual(values = c("solid", "twodash")) +

theme(legend.title = element_blank(),

legend.position = c(0.2, 0.2))

```

\# OP fig

```{r}

#op_virtual_fig <-

op_attendances %>%

filter(date_month >= ymd("2019-01-01")) %>%

filter(date_month < ymd("2021-01-01")) %>%

mutate(n_virtual = ifelse(n_virtual < 5, NA_real_, n_virtual)) %>%

pivot_longer(cols = -date_month,

names_to = "attendance_type",

values_to = "attendances") %>%

filter(attendance_type %in% c("n_attendances", "n_virtual")) %>%

filter(attendances >= 5) %>%

ggplot(aes(x = date_month, y = attendances, color = attendance_type)) +

geom_line(size = 1) +

scale_x_date(date_breaks = "5 month", date_labels = "%b %Y") +

labs(x = "", y = "Out-patient\n attendances per month\n") +

geom_vline(xintercept = as_date("2020-03-26"), color = "grey") +

geom_vline(xintercept = as_date("2020-07-21"), color = "grey") +

scale_color_manual(values = c("black", "dark orange"), labels = c("Total Outpatient", "Virtual Visit")) +

theme(legend.title = element_blank(), legend.position = "bottom", axis.text.x = element_text(size = 10))

```

#Clinics fig

```{r, fig.height=6, fig.width=7}

#op_clinics_fig <-

op_clinics %>%

filter(date_month < ymd("2020-08-01")) %>%

filter(date_month >= ymd("2019-01-01")) %>%

ggplot(aes(x = date_month, y = n)) +

geom_line() +

geom_vline(xintercept = as_date("2020-03-26"), color = "grey") +

geom_vline(xintercept = as_date("2020-07-31"), color = "grey") +

labs(x = "", y = "") +

facet_wrap(

~ fct_rev(fct_reorder(specialty_national, n, max)),

ncol = 3,

scales = "free",

labeller = label_wrap_gen()) +

expand_limits(y = 0) +

theme(legend.title = element_blank(),

axis.text = element_text(size = 8),

strip.text = element_text(size = 10),

strip.background = element_rect(fill = "white"))

```

#AE fig

```{r}

ae_admissions %>%

#filter(date_month <= ymd("2020-07-01")) %>%

ggplot(aes(x = date_month, y = total)) +

geom_vline(xintercept = as_date("2020-03-26"), color = "grey") +

geom_vline(xintercept = as_date("2020-06-18"), color = "grey") +

geom_line(size = 1, color = "black") +

ylim(0, 1000) +

labs(x = "", y = "A&E attendances / month")

```

#Tables

#Demog Table Function

```{r}

#create summary table from dataframe

create_summary_table <-

function(x) {

x %>%

summarise(

n_people = n(),

prop_shielding = n_people/n_shielders,

n_women = sum(sex == "F"),

prop_women = n_women / n_people,

median_age = median(age),

under_20 = sum(age < 20),

prop_children = under_20 / n_people,

median_simd = median(simd_decile, na.rm = T),

simd_1_or_2 = sum(simd_decile == 1 | simd_decile == 2, na.rm = T),

prop_deprived = simd_1_or_2 / n_people,

remote_or_rural = sum(ur_class %in% c(4:6)),

prop_remote_rural = remote_or_rural / n_people,

n_national = sum(source == "National"),

prop_national = n_national/n_people,

prop_multimorbid = sum(score > 1, na.rm = T) / n_people,

mean_wscore_vw = mean(wscore_vw, na.rm = T),

.groups = "drop") %>%

mutate(across(starts_with("prop"), round, 2))

}

```

#Demographics tables

```{r}

demog %>%

create_summary_table() %>%

write_csv("demog_totals.csv")

demog %>%

group_by(age_band) %>%

create_summary_table() %>%

write_csv("demog_age.csv")

demog %>%

group_by(sex) %>%

create_summary_table() %>%

write_csv("demog_sex.csv")

demog %>%

group_by(dominant_reason) %>%

create_summary_table() %>%

write_csv("demog_reason.csv")

demog %>%

group_by(source) %>%

create_summary_table() %>%

write_csv("demog_source.csv")

demog %>%

group_by(simd_decile) %>%

create_summary_table() %>%

write_csv("demog_simd.csv")

demog %>%

group_by(ur_name) %>%

create_summary_table() %>%

write_csv("demog_ur.csv")

```

#Precovid Care Summary Function

```{r}

#create summary table from dataframe

create_precovid_table <-

function(x) {

x %>%

summarise(

n_people = n(),

prop_shielding = n_people/n_shielders,

prop_op = sum(n_op_1yr > 0) / n_people,

prop_ip_scheduled = sum(n_routine_1yr > 0) / n_people,

prop_ip_emergency = sum(n_emerg_1yr > 0) / n_people,

prop_ae = sum(n_ae_1yr > 0) / n_people,

n_all_routine = sum(all_routine_1yr),

mean_routine = mean(all_routine_1yr),

n_all_emergency = sum(all_emergency_1yr),

mean_emergency = mean(all_emergency_1yr),

prop_emergency = n_all_emergency / (n_all_emergency + n_all_routine),

los_year = sum(los_smr01_1yr),

mean_los = mean(los_smr01_1yr),

prop_multimorbid = sum(score > 1, na.rm = T) / n_people,

mean_wscore_vw = mean(wscore_vw, na.rm = T),

.groups = "drop") %>%

mutate(across(starts_with("prop"), round, 2)) %>%

mutate(across(contains("mean"), round, 1))

}

```

#Pre-Covid care tables

```{r}

demog %>%

create_precovid_table() %>%

write_csv("precovid_totals.csv")

demog %>%

group_by(age_band) %>%

create_precovid_table() %>%

write_csv("precovid_age.csv")

demog %>%

group_by(sex) %>%

create_precovid_table() %>%

write_csv("precovid_sex.csv")

demog %>%

group_by(dominant_reason) %>%

create_precovid_table() %>%

write_csv("precovid_reason.csv")

demog %>%

group_by(source) %>%

create_precovid_table() %>%

write_csv("precovid_source.csv")

demog %>%

group_by(simd_decile) %>%

create_precovid_table() %>%

write_csv("precovid_simd.csv")

demog %>%

group_by(ur_name) %>%

create_precovid_table() %>%

write_csv("precovid_ur.csv")

```

#Lockdown summaries

```{r}

create_lockdown_usage_table <-

function(x) {

x %>%

summarise(

n_people = n(),

prop_op = sum(n_lockdown_op > 0) / n_people,

prop_smr01_routine = sum(n_lockdown_admissions_routine > 0) / n_people,

prop_smr01_emerg = sum(n_lockdown_admissions_emerg > 0) / n_people,

prop_ae = sum(n_lockdown_ae > 0) / n_people,

mean_op = mean(n_lockdown_op) * 100,

mean_smr01_routine = mean(n_lockdown_admissions_routine) * 100,

mean_smr01_emerg = mean(n_lockdown_admissions_emerg) * 100,

mean_ae = mean(n_lockdown_ae) * 100,

n_op = sum(n_lockdown_op),

n_smr01_routine = sum(n_lockdown_admissions_routine),

n_smr01_emerg = sum(n_lockdown_admissions_emerg),

n_ae = sum(n_lockdown_ae),

n_total_routine = n_op + n_smr01_routine,

n_total_emergency = n_ae + n_smr01_emerg,

prop_emergency = n_total_emergency / (n_total_routine + n_total_emergency),

n_dead = sum(dead == 1),

dead_per_k = round(n_dead / n_people * 1000, 0),

.groups = "drop") %>%

mutate(across(contains("mean"), round, 0)) %>%

mutate(across(contains("prop"), round, 2))

}

```

#Who lockdown care

```{r}

demog %>%

group_by(n_lockdown_ae > 0) %>%

create_summary_table() %>%

write_csv(., "lockdown_usage_ae.csv")

demog %>%

group_by(n_lockdown_ae_major > 0) %>%

create_summary_table() %>%

write_csv(., "lockdown_usage_ae_major.csv")

demog %>%

group_by(n_lockdown_admissions > 0) %>%

create_summary_table() %>%

write_csv(., "lockdown_usage_smr01.csv")

demog %>%

group_by(n_lockdown_admissions_emerg > 0) %>%

create_summary_table() %>%

write_csv(., "lockdown_usage_smr01_emerg.csv")

demog %>%

group_by(n_lockdown_admissions_routine > 0) %>%

create_summary_table() %>%

write_csv(., "lockdown_usage_smr01_routine.csv")

demog %>%

group_by(n_lockdown_op > 0) %>%

create_summary_table() %>%

write_csv(., "lockdown_usage_op.csv")

demog %>%

group_by(n_lockdown_op_virtual > 0) %>%

create_summary_table() %>%

write_csv(., "lockdown_usage_op_virtual.csv")

demog %>%

group_by(n_lockdown_op_return > 0) %>%

create_summary_table() %>%

write_csv(., "lockdown_usage_op_return.csv")

demog %>%

group_by(n_lockdown_op_new > 0) %>%

create_summary_table() %>%

write_csv(., "lockdown_usage_op_new.csv")

demog %>%

group_by(n_lockdown_op_adhoc > 0) %>%

create_summary_table() %>%

write_csv(., "lockdown_usage_op_adhoc.csv")

demog %>%

group_by(no_visits = n_lockdown_admissions == 0 & n_lockdown_op == 0 & n_lockdown_ae == 0) %>%

create_summary_table() %>%

write_csv(., "lockdown_usage_no_visits.csv")

```

#What lockdown care

```{r}

demog %>%

create_lockdown_usage_table() %>%

write_csv(., "lockdown_usage_total.csv")

demog %>%

group_by(source) %>%

create_lockdown_usage_table() %>%

write_csv(., "lockdown_usage_source.csv")

demog %>%

group_by(sex) %>%

create_lockdown_usage_table() %>%

write_csv(., "lockdown_usage_sex.csv")

demog %>%

group_by(age_band) %>%

create_lockdown_usage_table() %>%

write_csv(., "lockdown_usage_age.csv")

demog %>%

group_by(simd_decile) %>%

create_lockdown_usage_table() %>%

write_csv(., "lockdown_usage_simd.csv")

demog %>%

group_by(dominant_reason) %>%

create_lockdown_usage_table() %>%

write_csv(., "lockdown_usage_reason.csv")

demog %>%

group_by(ur_name) %>%

create_lockdown_usage_table() %>%

write_csv(., "lockdown_usage_ur.csv")

demog %>%

group_by(index) %>%

create_lockdown_usage_table() %>%

write_csv(., "lockdown_usage_mm.csv")

```

\# Health Care Resource Utilisation (HCRU) by Grampian vs Shielding populations

\# Generalised Additive Model using mgcv library

#_______________________________________________________________________________

\# library ----

library(data.table)

library(bit64)

library(mgcv)

#_______________________________________________________________________________

\# Data ----

\# Data start date, lockdown start date and lockdown end date

dsDate <- as.IDate('2020-01-12')

lsDate <- as.IDate('2020-03-15')

leDate <- as.IDate('2020-06-18')

exclDate <- as.IDate('2020-03-15')

#_______________________________________________________________________________

\# Fn to organise data -----

fn_createData <- function(oDT){

\# Subset data

wDT <- oDT[date_week <= leDate,]

\# Reshape data

setcolorder(wDT, c('date_week',

'grampian_total', 'shielding_total',

'grampian_total_per_k', 'shielding_total_per_k'))

value_name <- c('total', 'total_per_k')

DT <- melt(wDT, id.vars = 'date_week',

measure.vars = patterns('.*total$', 'total_per_k'),

variable.name = 'Group',

value.name = value_name)

\# Group variable

DT$Group <- as.character(DT$Group)

DT[, Group := ifelse(Group == '1', 'G', 'S')]

\# Create Phase variable

DT$Phase <- NA_character_

DT[, Phase := ifelse(date_week <= lsDate, 'P1', 'P2')]

\# str(DT)

\# Group_Phase

DT[, Group_Phase := as.factor(paste0(Group, '_', Phase))]

\# Week

DT[, iWeek := as.integer((date_week - dsDate) %/% 7 + 1)]

\# Integer N

DT[, N := as.integer(total)]

DT[Group == 'G', NP := 585700]

DT[Group == 'S', NP := 16092]

\# Remove Week 10

DT <- DT[!(iWeek == 10), ]

return(DT)

}

#_______________________________________________________________________________

\# Outpatient ----

\# ALL OPD (TRAK)

fname <- 'trak_outpatient_per_week_total.csv'

OPD <- fread(file = paste0(datDir, fname), sep = ',')

OPD$date_week <- as.IDate(OPD$date_week)

OPD <- OPD[date_week != exclDate & date_week >= dsDate & date_week <= leDate,]

DT <- fn_createData(OPD)

\# FINAL MODEL

fm_NB <- mgcv::gam(N ~ Group_Phase + iWeek + offset(log(NP)),

family = nb(link = 'log'),

method = 'REML', gamma = 1,

na.action = na.omit,

data = DT[!(iWeek == 10), ])

summary(fm_NB)

anova(fm_NB)

#_______________________________________________________________________________

#_______________________________________________________________________________

\# Inpatient Non-Emergency (Scheduled) ----

\# Data

fname <- 'smr01_inpatient_per_week_total.csv'

IP <- fread(file = paste0(datDir, fname), sep = ',')

IP$date_week <- as.IDate(IP$date_week)

IP <- IP[date_week != exclDate & date_week >= dsDate & date_week <= leDate,]

IP <- IP[admission_type == 'scheduled', ]

DT <- fn_createData(IP)

\# FINAL MODEL

fm_NB <- mgcv::gam(N ~ Group_Phase + iWeek + offset(log(NP)),

family = nb(link = 'log'),

method = 'REML', gamma = 1,

na.action = na.omit,

data = DT[!(iWeek == 10), ])

summary(fm_NB)

anova(fm_NB)

#_______________________________________________________________________________

#_______________________________________________________________________________

\# Inpatient Emergency ----

\# Data

fname <- 'smr01_inpatient_per_week_total.csv'

IP <- fread(file = paste0(datDir, fname), sep = ',')

IP$date_week <- as.IDate(IP$date_week)

IP <- IP[date_week != exclDate & date_week >= dsDate & date_week <= leDate,]

IP <- IP[admission_type == 'emergency', ]

DT <- fn_createData(IP)

\# FINAL MODEL

fm_NB <- mgcv::gam(N ~ Group_Phase + iWeek + offset(log(NP)),

family = nb(link = 'log'),

method = 'REML', gamma = 1,

na.action = na.omit,

data = DT[!(iWeek == 10), ])

summary(fm_NB)

anova(fm_NB)

#_______________________________________________________________________________

#_______________________________________________________________________________

\# Accident & Emergency -----

\# ALL AE (TRAK)

fname <- 'trak_emergency_per_week_total.csv'

AE <- fread(file = paste0(datDir, fname), sep = ',')

AE$date_week <- as.IDate(AE$date_week)

AE <- AE[date_week != exclDate & date_week >= dsDate & date_week <= leDate,]

DT <- fn_createData(AE)

\# FINAL MODEL

fm_NB <- mgcv::gam(N ~ Group_Phase + iWeek,

family = nb(link = 'log'),

method = 'REML', gamma = 1,

na.action = na.omit, nthreads = nt,

data = DT[!(iWeek == 10), ])

summary(fm_NB)

anova(fm_NB)

#_______________________________________________________________________________

\# Fitting GLMM on summarised data of Shielding patients

\# Uses summarised data for the combination of: (Phase, sex, AGEgr_, SIMDgr_, SHgroup_)

\# Fit using glmmTMB library

#_______________________________________________________________________________

\# library ----

library(data.table)

library(glmmTMB)

library(parallel)

#_______________________________________________________________________________

#_______________________________________________________________________________

\# OPD ----

\# OPD

load('S_OPD.RData')

\# Data summary

DT <- fDT[, .(nEvent = sum(N), nPat = .N, iDay = median(iDay)),

by = .(Phase, sex, AGEgr_, SIMDgr_, SHgroup_)]

DT$gID <- as.factor(paste0('R', rep(1:(nrow(DT)/2), each = 2)))

\# Intermediate Model (NB)

fm_NB <- glmmTMB::glmmTMB(nEvent ~ sex + SIMDgr_ + AGEgr_ + SHgroup_ + Phase +

sex:Phase + SIMDgr_:Phase + AGEgr_:Phase + SHgroup_:Phase +

offset(log(iDay)) + offset(log(nPat)) + (1 | gID),

family = nbinom2(link = 'log'),

data = DT,

control = glmmTMBControl(parallel = parallel::detectCores()))

summary(fm_NB)

fm <- fm_NB

save(fm, file = 'fm_OPD_NB_int.RData')

\# Final Model (NB)

fm_NB <- glmmTMB::glmmTMB(nEvent ~ sex + SIMDgr_ + AGEgr_ + SHgroup_ + Phase +

sex:Phase + AGEgr_:Phase + SHgroup_:Phase +

AGEgr_:SHgroup_ + AGEgr_:SHgroup_:Phase +

offset(log(iDay)) + offset(log(nPat)) + (1 | gID),

family = nbinom2(link = 'log'),

data = DT,

control = glmmTMBControl(parallel = parallel::detectCores()))

summary(fm_NB)

fm <- fm_NB

save(fm, file = 'fm_OPD_NB_final.RData')

#_______________________________________________________________________________

#_______________________________________________________________________________

\# IP Non-Emergency (Scheduled) ----

\# IP

load('S_IP.RData')

fDT$N <- fDT$N_NonEmerg

\# Data summary

DT <- fDT[, .(nEvent = sum(N), nPat = .N, iDay = median(iDay)),

by = .(Phase, sex, AGEgr_, SIMDgr_, SHgroup_)]

DT$gID <- as.factor(paste0('R', rep(1:(nrow(DT)/2), each = 2)))

\# Intermediate Model (NB)

fm_NB <- glmmTMB::glmmTMB(nEvent ~ sex + SIMDgr_ + AGEgr_ + SHgroup_ + Phase +

sex:Phase + SIMDgr_:Phase + AGEgr_:Phase + SHgroup_:Phase +

offset(log(iDay)) + offset(log(nPat)) + (1 | gID),

family = nbinom2(link = 'log'),

data = DT,

control = glmmTMBControl(parallel = parallel::detectCores()))

summary(fm_NB)

fm <- fm_NB

save(fm, file = 'fm_IP_NonEmerg_NB_int.RData')

\# Final model (NB)

fm_NB <- glmmTMB::glmmTMB(nEvent ~ sex + SIMDgr_ + AGEgr_ + SHgroup_ + Phase +

AGEgr_:Phase + SHgroup_:Phase +

offset(log(iDay)) + offset(log(nPat)) + (1 | gID),

family = nbinom2(link = 'log'),

data = DT,

control = glmmTMBControl(parallel = parallel::detectCores()))

summary(fm_NB)

fm <- fm_NB

save(fm, file = 'fm_IP_NonEmerg_NB_final.RData')

#_______________________________________________________________________________

#_______________________________________________________________________________

\# IP Emergency----

\# IP

load('S_IP.RData')

fDT$N <- fDT$N_Emerg

\# Data summary

DT <- fDT[, .(nEvent = sum(N), nPat = .N, iDay = median(iDay)),

by = .(Phase, sex, AGEgr_, SIMDgr_, SHgroup_)]

DT$gID <- as.factor(paste0('R', rep(1:(nrow(DT)/2), each = 2)))

\# Intermediate Model (NB)

fm_NB <- glmmTMB::glmmTMB(nEvent ~ sex + SIMDgr_ + AGEgr_ + SHgroup_ + Phase +

sex:Phase + SIMDgr_:Phase + AGEgr_:Phase + SHgroup_:Phase +

offset(log(iDay)) + offset(log(nPat)) + (1 | gID),

family = nbinom2(link = 'log'),

data = DT,

control = glmmTMBControl(parallel = parallel::detectCores()))

summary(fm_NB)

fm <- fm_NB

save(fm, file = 'fm_IP_Emerg_NB_int.RData')

\# Final model (NB)

fm_NB <- glmmTMB::glmmTMB(nEvent ~ sex + SIMDgr_ + AGEgr_ + SHgroup_ + Phase +

offset(log(iDay)) + offset(log(nPat)) + (1 | gID),

family = nbinom1(link = 'log'),

data = DT,

control = glmmTMBControl(parallel = parallel::detectCores()))

summary(fm_NB)

fm <- fm_NB

save(fm, file = 'fm_IP_Emerg_NB_final.RData')

#_______________________________________________________________________________

#_______________________________________________________________________________

\# Accident & Emergency ----

\# AE

load('S_AE.RData')

\# Data summary

DT <- fDT[, .(nEvent = sum(N), nPat = .N, iDay = median(iDay)),

by = .(Phase, sex, AGEgr_, SIMDgr_, SHgroup_)]

DT$gID <- as.factor(paste0('R', rep(1:(nrow(DT)/2), each = 2)))

\# Intermediate Model (NB)

fm_NB <- glmmTMB::glmmTMB(nEvent ~ sex + SIMDgr_ + AGEgr_ + SHgroup_ + Phase +

sex:Phase + SIMDgr_:Phase + AGEgr_:Phase + SHgroup_:Phase +

offset(log(iDay)) + offset(log(nPat)) + (1 | gID),

family = nbinom2(link = 'log'),

data = DT,

control = glmmTMBControl(parallel = parallel::detectCores()))

summary(fm_NB)

fm <- fm_NB

save(fm, file = 'fm_AE_NB_int.RData')

\# Final model (NB)

fm_NB <- glmmTMB::glmmTMB(nEvent ~ sex + SIMDgr_ + AGEgr_ + SHgroup_ + Phase +

AGEgr_:Phase + AGEgr_:SHgroup_ +

offset(log(iDay)) + offset(log(nPat)) + (1 | gID),

family = nbinom1(link = 'log'),

data = DT,

control = glmmTMBControl(parallel = parallel::detectCores()))

summary(fm_NB)

fm <- fm_NB

save(fm, file = 'fm_AE_NB_final.RData')

#_______________________________________________________________________________
