## Supplementary Figure for "The clinically extremely vulnerable to COVID: Identification and changes in healthcare while self-isolating (shielding) during the coronavirus pandemic"

**Supplementary Figure 1. Comparison of mean visits pre-lockdown and during lockdown by age and reason for shielding**. Pre-lockdown period is 1 January to 14 March 2020 in black and the lockdown period is 22 March to 18 June 2020 in red. *Note the Y axis scale varies for each care type*.

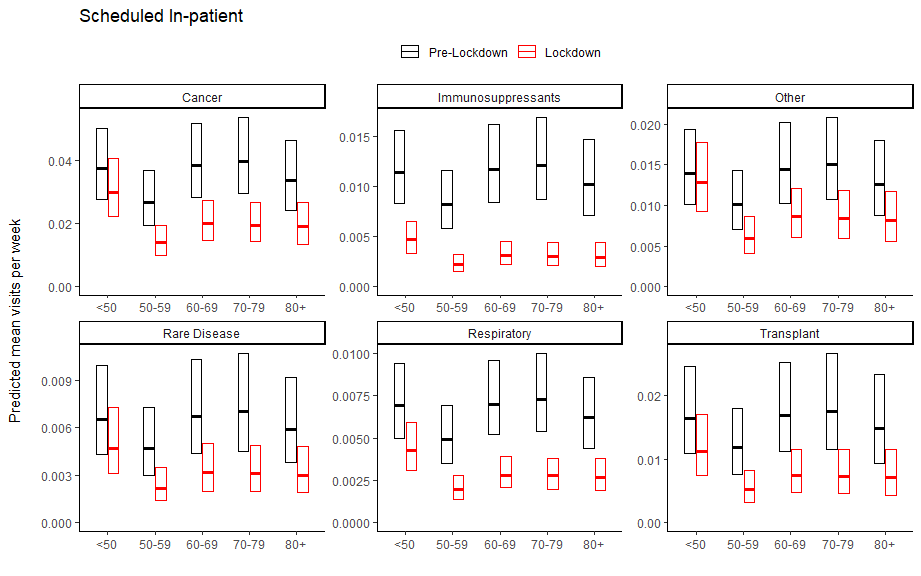

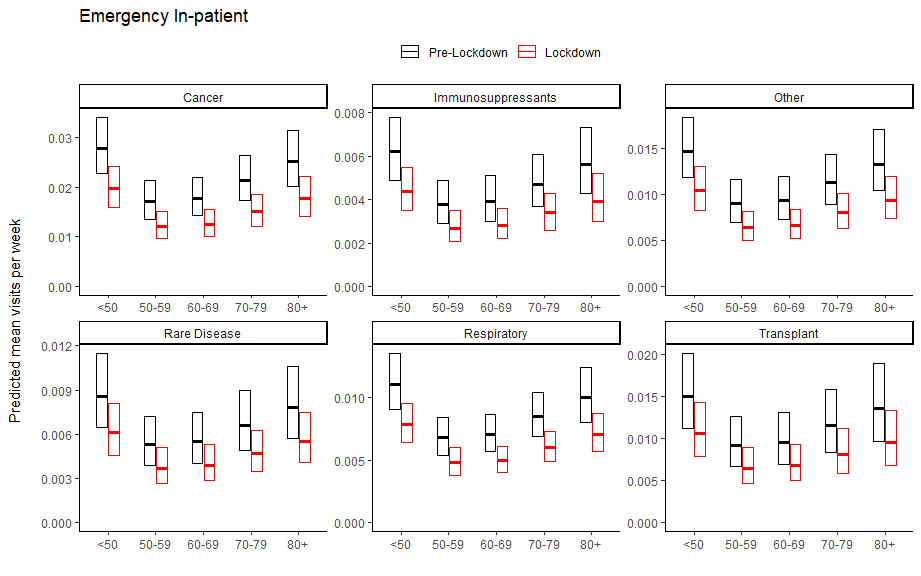

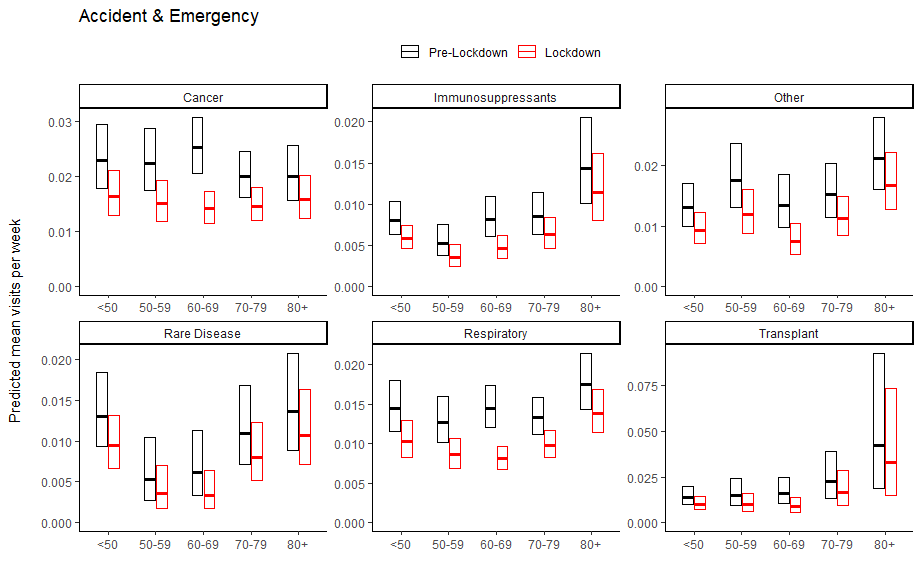
